# Arrhythmia variant associations and reclassifications in the eMERGE-III sequencing study

**DOI:** 10.1101/2021.03.30.21254549

**Authors:** Andrew M. Glazer, Giovanni Davogustto, Christian M. Shaffer, Carlos G. Vanoye, Reshma R. Desai, Eric H. Farber-Eger, Ozan Dikilitas, Ning Shang, Jennifer A. Pacheco, Tao Yang, Ayesha Muhammad, Jonathan D. Mosley, Sara L. Van Driest, Quinn S. Wells, Lauren Lee Rinke, Olivia R. Kalash, Yuko Wada, Sarah Bland, Zachary T. Yoneda, Devyn W. Mitchell, Brett M. Kroncke, Iftikhar J. Kullo, Gail P. Jarvik, Adam S. Gordon, Eric B. Larson, Teri A. Manolio, Tooraj Mirshahi, Jonathan Z. Luo, Daniel Schaid, Bahram Namjou, Tarek Alsaied, Rajbir Singh, Ashutosh Singhal, Cong Liu, Chunhua Weng, George Hripcsak, James D. Ralston, Elizabeth M. McNally, Wendy K. Chung, David S. Carrell, Kathleen A. Leppig, Hakon Hakonarson, Patrick Sleiman, Sunghwan Sohn, Joseph Glessner, the eMERGE Network, Joshua Denny, Wei-Qi Wei, Alfred L. George, M. Benjamin Shoemaker, Dan M. Roden

**Affiliations:** Vanderbilt University Medical Center, Nashville TN; Northwestern University, Chicago IL; Mayo Clinic, Rochester MN; Columbia University Irving Medical Center, New York NY; Departments of Medicine (Medical Genetics) and Genome Sciences, University of Washington School of Medicine, Seattle, WA; Kaiser Permanente of Washington, Seattle, WA; National Human Genome Research Institute, NIH, Bethesda MD; Geisinger Health System, Danville PA; Cincinnati Children’s Hospital Medical Center, Cincinnati OH; Meharry Medical College, Nashville TN; Children’s Hospital of Philadelphia, Philadelphia PA; All of Us Research Program, NIH, Bethesda MD

## Abstract

In 21,846 eMERGE-III participants, sequencing 10 arrhythmia syndrome disease genes identified 123 individuals with pathogenic or likely pathogenic (P/LP) variants. Compared to non-carriers, P/LP carriers had a significantly higher burden of arrhythmia phenotypes in their electronic health records (EHRs). Fifty one participants had variant results returned. Eighteen of these 51 participants had inherited arrhythmia syndrome diagnoses (primarily long QT syndrome), and 11/18 of these diagnoses were made only after variant results were returned. After *in vitro* functional evaluation of 50 variants of uncertain significance (VUS), we reclassified 11 variants: 3 to likely benign and 8 to P/LP. As large numbers of people are sequenced, the disease risk from rare variants in arrhythmia genes can be assessed by integrating genomic screening, EHR phenotypes, and *in vitro* functional studies.

## Introduction

The increasing use of large-scale sequencing and genomic medicine holds great promise for identifying individuals at risk for rare Mendelian diseases. The American College of Genetics and Genomics (ACMG) has recommended that incidentally discovered pathogenic/likely pathogenic (P/LP) variants in 59 Mendelian disease genes be reported back to patients.^1^ However, two major challenges remain to implementation of broad scale genomic screening. First, P/LP variants often exhibit incomplete penetrance,^2,3^ so it is unclear what fraction of individuals with incidentally discovered P/LP variants will have a phenotype. Second, the vast majority of coding variants in Mendelian disease genes are currently classified as variants of uncertain significance (VUS).^4,5^ Common approaches to VUS reclassification have involved segregation studies in individual families or *in vitro* functional experiments.^6-9^

Congenital arrhythmia syndromes are rare Mendelian diseases that can cause sudden unexplained death in otherwise healthy individuals. These syndromes, which include long QT syndrome (LQTS), Brugada syndrome (BrS), *LMNA*-related dilated cardiomyopathy and arrhythmia, and Catecholaminergic Polymorphic Ventricular Tachycardia (CPVT), affect approximately 1 in 1,000 to 1 in 10,000 individuals.^10-13^ These syndromes are typically caused by dominant rare pathogenic variants in cardiac ion channel genes and other related genes.^14^ Individuals with these disorders have presentations including diverse arrhythmias and electrocardiographic abnormalities, syncope, cardiomyopathy, or sudden death. It is standard of care to provide presymptomatic genetic screening and prophylactic therapy (if indicated) to relatives of affected probands.^15^ However, the role of genomic screening across populations for these syndromes is not yet established.

The multicenter Electronic Medical Records and Genomics sequencing study (eMERGE-III) investigated the feasibility of genomic screening by sequencing 109 genes implicated across the spectrum of Mendelian diseases in over 20,000 individuals, returning variant results to the participants, and using Electronic Health Record (EHR) and follow-up clinical data to ascertain patient phenotypes.^16,17^ In this work, we investigate the association between rare variants in 10 arrhythmia genes with arrhythmia phenotypes in 21,846 eMERGE-III participants unselected for arrhythmia phenotypes. In addition, we assess the utility of using genomic screening, EHR phenotypes, and *in vitro* functional datasets to reclassify VUS in arrhythmia genes.

## Results

### Patient characteristics and genotypes

We studied 21,846 eMERGE-III participants who did not have a clinical indication for sequencing related to arrhythmias, cardiomyopathy, or heart failure. Some participants had clinical diagnoses related to other conditions (e.g. colon cancer).^17^ The median age of the study group was 58 (IQR: 26-70), 10.9% were under 18 years old at the time of record review, 52.2% were female, and 69.8% self-reported as White (Table S1). We focused on 10 genes previously linked to Mendelian arrhythmia syndromes (*ANK2, CACNA1C, KCNE1, KCNE2, KCNH2, KCNJ2, KCNQ1, LMNA, RYR2*, and *SCN5A*; Table S2).^16^ Among these genes, 123 individuals (0.56%) were heterozygous for a P/LP variant according to American College of Genetics and Genomics/American College of Pathology (ACMG/AMP) criteria.^18^ In addition, 1,838 individuals (8.4%) were not heterozygous for any P/LP variant, but were heterozygous for one or more of 1,648 ultra-rare VUS (gnomAD allele frequency below 2.5e-5;^19,20^ Table S1). Self-reported White and non-White individuals had a similar frequency of P/LP variants (0.53% for Whites vs. 0.65% for non-Whites, p=0.28). However, compared to White individuals, non-Whites were more likely to be heterozygous for rare VUS (7.4% for Whites vs. 10.8% for non-Whites, p=3.8e-19).

### Association between pathogenic variants and EHR-derived phenotypes

Compared to individuals without any P/LP variants, participants heterozygous for P/LP variants had a higher prevalence of EHR code-based arrhythmia phenotypes (Figure 1, Table S3). These were concentrated in “extreme” and “severe” arrhythmia groups defined in the Methods section. For example, 22/123 (18%) of all P/LP carriers had an EHR-documented extreme or severe arrhythmia code compared to 1,386/19,885 (7%) of non-carriers (Figure 1). Using logistic regressions, we detected 15 significant associations with a false discovery rate (FDR) below 0.1 between arrhythmia EHR diagnoses and P/LP variants in all pooled genes or in specific genes, with odds ratios from 2.7 to 36.8 (Table S3). The most frequently associated disease was LQTS, which was associated with P/LP variants in all genes in a pooled analysis, as well as P/LP variants in *KCNE1, KCNH2*, and *KCNQ1* individually (Figure 2). P/LP variants in *LMNA* were associated with 7 of the 11 analyzed arrhythmia phenotypes, highlighting the pleiotropic nature of *LMNA*-related cardiomyopathy and arrhythmia. In contrast to the P/LP associations, we detected almost no associations of VUS with arrhythmia phenotypes (Figure 1, Table S4). The only exception was *LMNA* VUS, which were significantly associated with pooled extreme and severe phenotypes (Figure 1). Burden tests using the Optimized Sequence Kernel Association Test method (SKAT-O) yielded similar results to the logistic regression analyses; 12 of the 15 associations that were significant (FDR<0.1) with the logistic regression analyses were also significant in the SKAT-O analyses (Table S5, S6).

**Figure 1:**
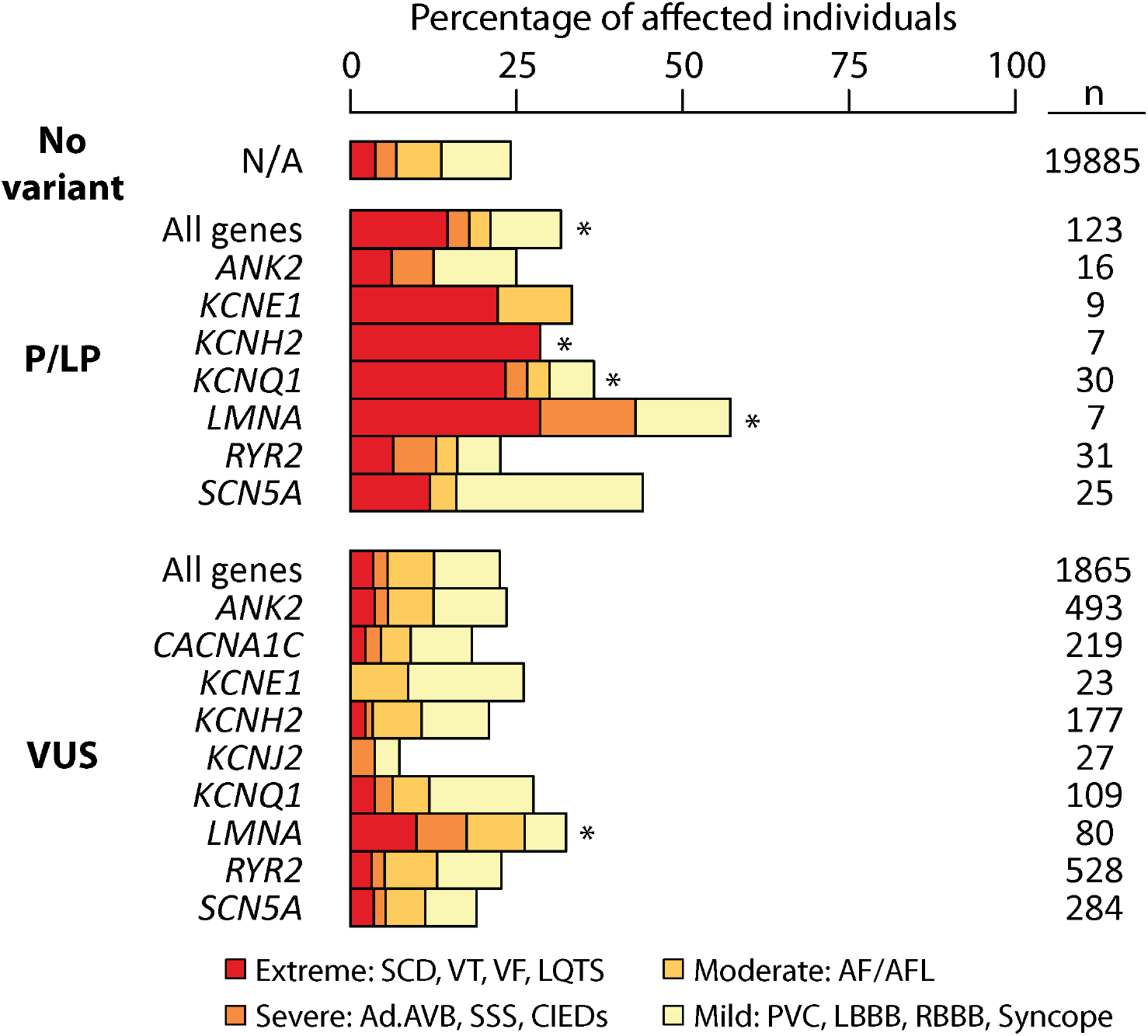
Penetrance of arrhythmia variants by gene. Abbreviations: SCD: sudden cardiac death, LQTS—long QT syndrome, VT—ventricular tachycardia, VF—ventricular fibrillation, AF/AFL—atrial fibrillation/atrial flutter, PVC— premature ventricular contraction, Ad.AVB—advanced atrioventricular block, SSS—sick sinus syndrome, CIEDs—cardiovascular implantable electronic devices, LBBB—left bundle branch block, RBBB—right bundle branch block, P/LP—pathogenic/likely pathogenic, VUS—variant of uncertain significance. * indicates false discovery rate < 0.1 (adjusted for the p-values from comparisons made in this figure). P values were derived from a logistic regression analysis which modeled severe or extreme phenotype status as a function of variant carrier status (P/LP vs. no variant or VUS vs. no variant), sex, site, age, and the first 10 principal components of ancestry.

**Figure 2:**
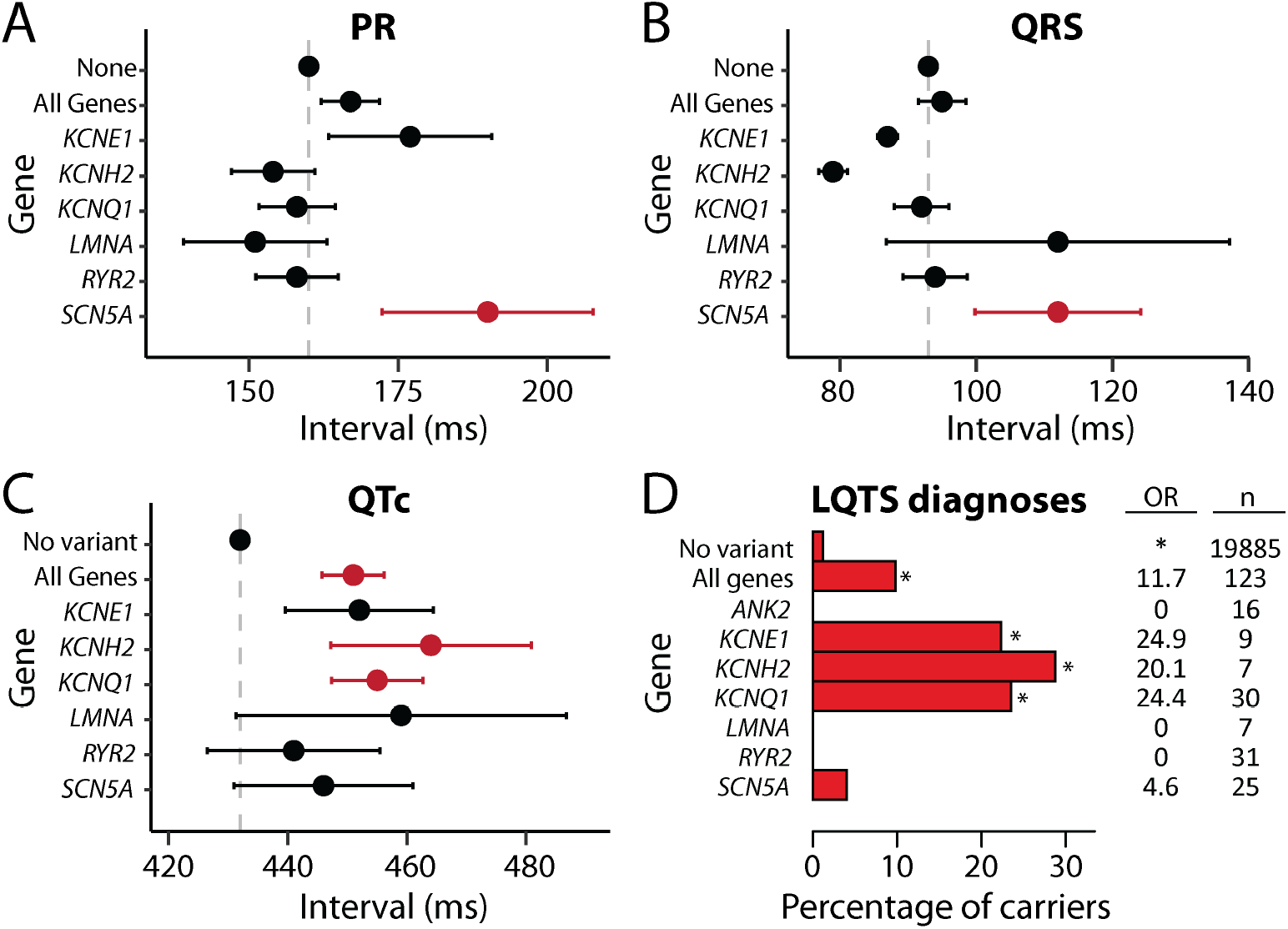
ECG interval associations and rate of LQTS diagnoses. A-D) Comparison between individuals with P/LP variants in all genes or in specific genes and individuals with no P/LP or ultra-rare VUS (“no variant”). Only groups with at least 5 carriers are shown. A-C) Electrocardiographic intervals (mean+/-SE) derived from the EHR from 4 of the 10 sites in the study. Red indicates significant associations (FDR<0.1), when compared to the no variant group. FDRs were calculated from all the p-values in panels A-C. D) Percentage of variant carriers with a long QT syndrome (LQTS) code in their EHR. OR=odds ratio, compared to individuals with no P/LP or ultra-rare VUS (“no variant”). *FDR<0.1. FDRs for panel D were calculated from all the p-values in Table S3 (including the other EHR-based phenotypes analyzed in this study). An additional summary of the data in panel D is presented in Table S9, including numbers of individuals in each group, and a similar analysis for VUS is presented in Table S10.

We performed a sensitivity analysis to determine if the code-based associations were simply due to a higher rate of arrhythmia diagnoses after 2017, when return of genetic results began, but this was not the case. Nearly all the associations remained intact when analyses were restricted to EHR codes before 2017, although some had a smaller effect size (Table S7). Similarly, when we restricted the analysis to the 70% of participants who self-reported as White, we observed concordant results, although again some of the associations had lower levels of significance (Table S8).

### Association between pathogenic variants and electrocardiographic parameters

To analyze the impact of genetic variants on electrocardiographic (ECG) intervals, we obtained all available ECGs from the EHR for 4 of the 10 sites (83,455 qualifying ECGs from 7,670 participants). Consistent with the established role of sodium current in regulating cardiac conduction velocity, we found significant associations between P/LP variants in *SCN5A* (encoding the cardiac sodium channel) and conduction delay (PR and QRS intervals; Figure 2A-B, Table S9). We also observed significant associations between the QTc interval and P/LP variants in all genes, and in individual potassium channel genes (*KCNQ1, KCNH2*, and *KCNE1;* Figure 2C). There was also directional concordance between the quantitative QTc interval effects (Figure 2C) and LQTS diagnosis associations (Figure 2D). There were no significant associations between VUS and ECG intervals (Table S10).

### Return of variant results

51 participants with P/LP variants in selected arrhythmia genes (*KCNE1, KCNH2, KCNQ1, SCN5A*) were returned variant results (RoR, Figure 3). All were encouraged to seek follow-up clinical care and screening. One year after receiving results, 28 of the 51 (55%) participants had a documented arrhythmia phenotype. 18 of the 51 (35%) received an inherited arrhythmia diagnosis (15 long QT syndrome, 2 Brugada syndrome, 1 *LMNA*-related DCM). Seven diagnoses were made before and 11 were made after RoR. These patients had a high prevalence of ECG abnormalities and other arrhythmias: 14 out of 18 participants had arrhythmia phenotypes or abnormal electrocardiographic phenotypes such as QTc> 480 ms or a Brugada ECG pattern (Table S11). The remaining 4 participants were diagnosed with an arrhythmia syndrome but had borderline phenotypes or no documented phenotypes. Changes in clinical management following RoR included: a new diagnosis (11), medication change (6), medication counseling (3), and Implantable Cardioverter Defibrillator placement (1) (Table S12).

**Figure 3:**
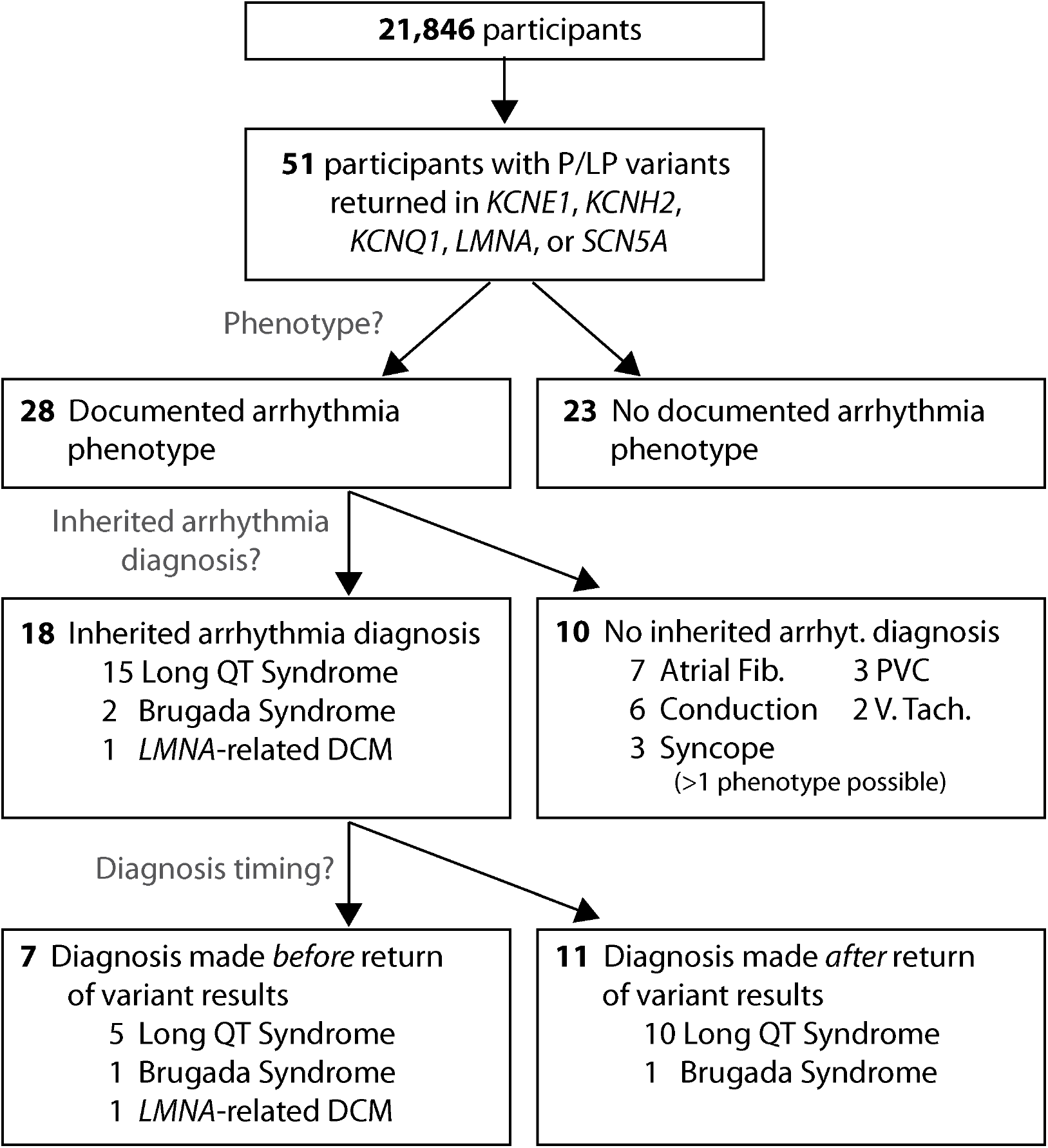
Return of variant results. Abbreviations: DCM—Dilated cardiomyopathy; Atrial Fib—Atrial Fibrillation; PVC— Premature Ventricular Contractions; V. Tach—Ventricular Tachycardia.

### VUS and variant reclassification

Ultra-rare VUS (gnomAD allele frequency <2.5e-5) were considered plausible candidate arrhythmia-associated variants.^20^ Indeed, 116 ultra-rare VUS occurred in at least 1 participant with an extreme or severe arrhythmia phenotype (Figure 4, Figure S1). It was not possible in this study to precisely determine the arrhythmia penetrance of ultra-rare variants since they were (by definition) present in only 1 or a few individuals. Nevertheless, to test the hypothesis that genomic screening and EHR-derived phenotypes could help identify disease-causing variants, we used heterologous expression and *in vitro* electrophysiological recording to evaluate the function of 52 ultra-rare eMERGE-III variants in *KCNQ1, KCNH2*, and *SCN5A* (Figure 5). The initial ACMG/AMP classifications of the variants were 50 VUS, 1 likely benign (LB), and 1 LP.

**Figure 4:**
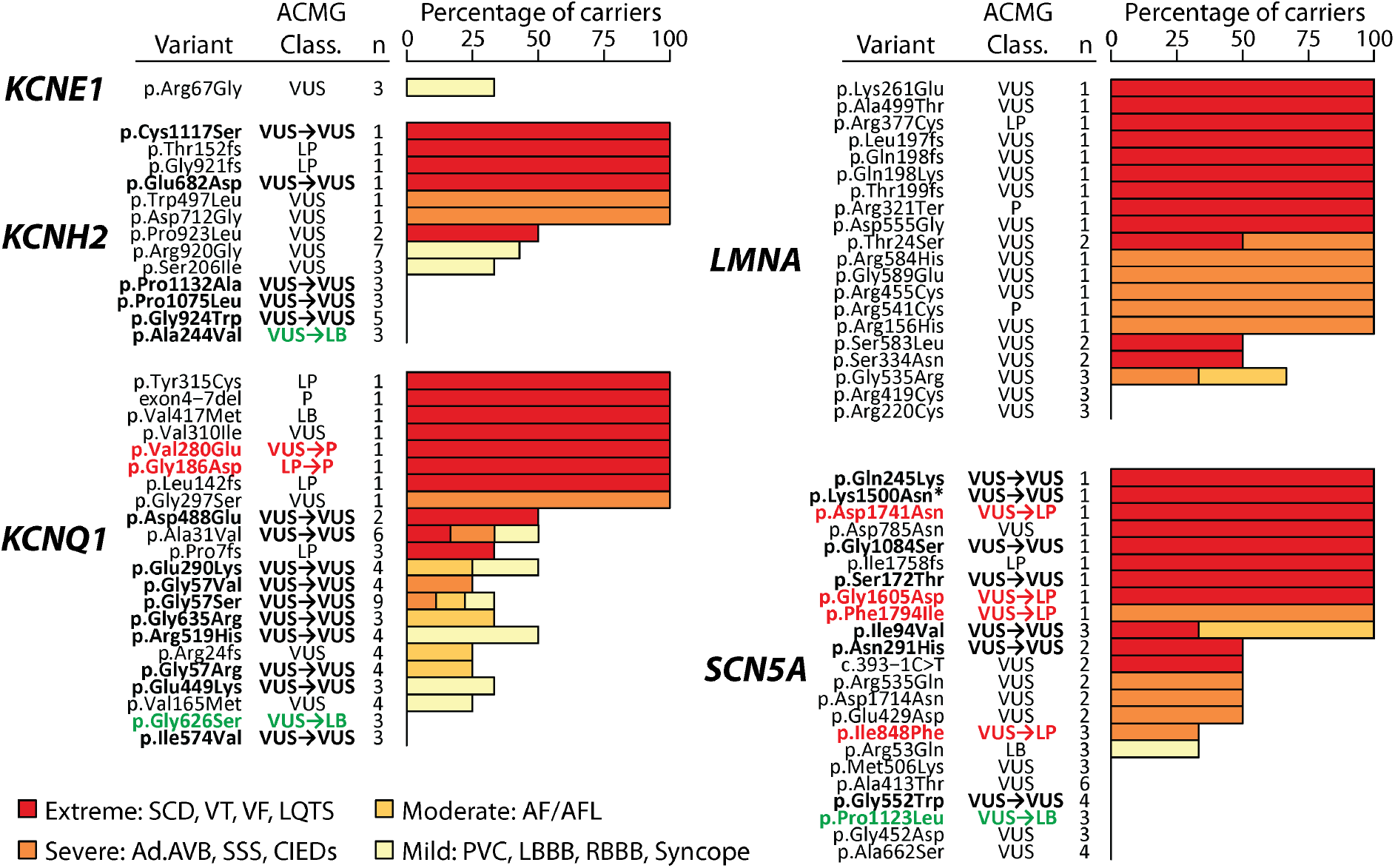
Percentage of variant carriers with arrhythmia phenotypes. Text columns indicate gene, variant name, variant ACMG classification, and number of individuals who were heterozygous for that variant in this study. Selected variants in *KCNE1, KCNH2, KCNQ1, LMNA*, and *SCN5A* are shown. A similar plot for the remaining genes in this study is presented in Figure S1. Shown variants have a gnomAD minor allele frequency below 2.5e-5 and are either 1) present in at least 3 eMERGE-III heterozygous participants or 2) present in at least 1 heterozyous participant with an extreme or severe phenotype. *Clinical genetic testing indicated that this participant also has a pathogenic variant in *DSP* p.Arg1269Ter and a VUS in *JUP* p.Ser323Gly. Variants in bold were subjected to *in vitro* functional studies (Table 1). Variant reclassifications are indicated with red or green text and are based on a combination of *in vitro* functional data and revised hotspot criteria (Table 1). Abbreviations: SCD—sudden cardiac death, LQTS—long QT syndrome, VT—ventricular tachycardia, VF—ventricular fibrillation, AF/ AFL—atrial fibrillation/atrial flutter, PVC—premature ventricular contraction, Ad.AVB—advanced atrioventricular block, SSS—sick sinus syndrome, CIEDs—cardiovascular implantable electronic devices, LBBB—left bundle branch block, RBBB—right bundle branch block, P/LP—pathogenic/likely pathogenic, VUS—variant of uncertain significance.

**Figure 5:**
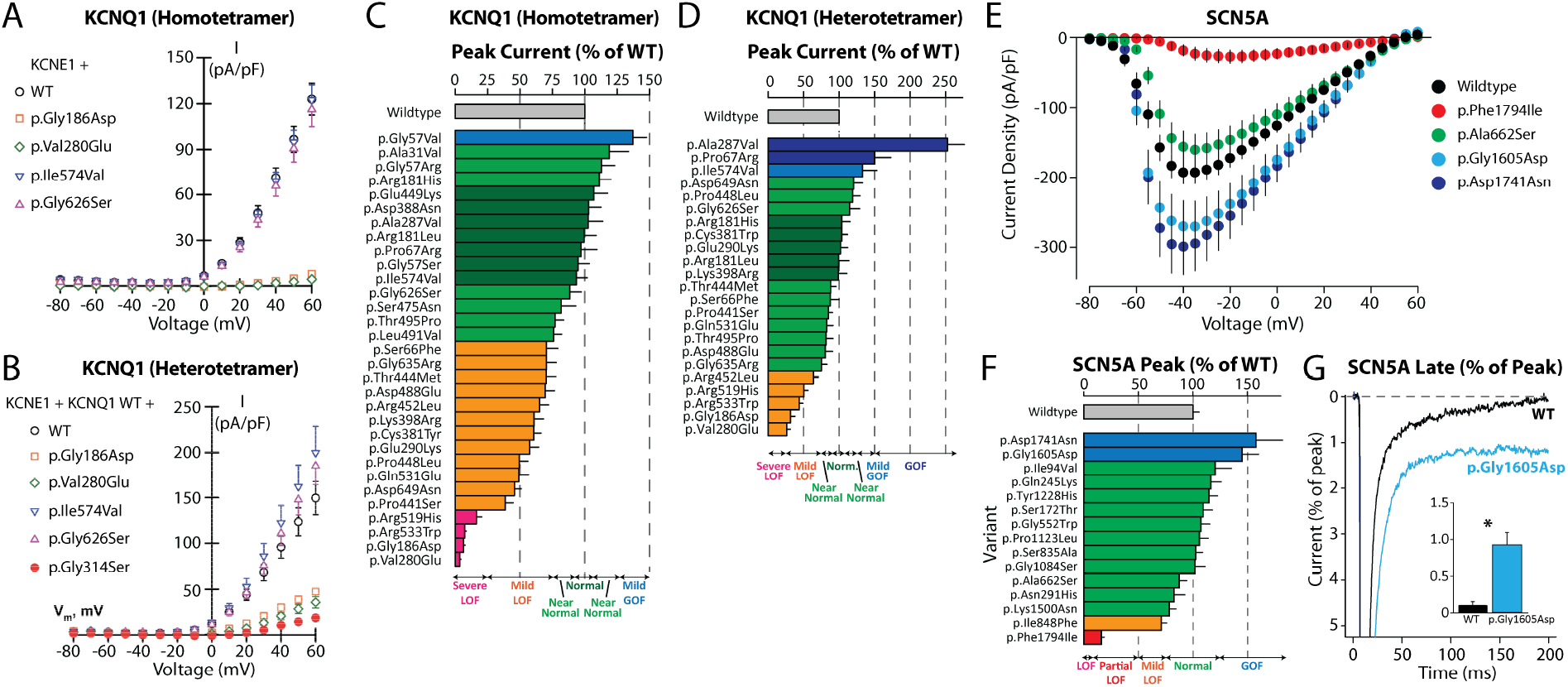
*In vitro* functional characterization of *KCNQ1* and *SCN5A* variants. A. Representative homotetramer current-voltage plots for selected *KCNQ1* variants. B. Representative heterotetramer current-voltage plots for selected *KCNQ1* variants. p.Gly314Ser is a control variant with dominant negative properties. For A-B wildtype *KCNE1* was coexpressed and for B a 1:1 ratio of WT:variant *KCNQ1* was expressed. C. *KCNQ1* current density for all variants studied in homotetrameric form. D. *KCNQ1* current density for all variants studied in heterotetrameric form. E. Representative current-voltage plots for selected *SCN5A* variants. F. *SCN5A* peak current density for all variants studied. Peak current density was measured as pA/pF and normalized to wildtype. G. Representative and quantification (inset) of late-current tracings for wildtype and p.Gly1605Asp variant. Late current measurements were calculated as a percentage of peak current from the same cell. *p=0.0028. Data in A-F were generated using the SyncroPatch instrument. Data in G were generated with manual patch-clamping. Chinese Hamster Ovary cells were used for A-D and Human Embryonic Kidney 293 cells were used for E-G. At least 20 replicate cells were analyzed per variant. The full patch clamp dataset, including numbers of replicate cells for each variant are listed in File S2.

Of the 52 studied variants, 28 had normal or near-normal *in vitro* function and were assigned the ACMG/AMP BS3 criterion. 5 variants had abnormal *in vitro* function strongly consistent with pathogenicity and were assigned PS3 at the strong level. 2 *KCNQ1* variants had severe loss of function as homotetramers but did not have strong dominant negative effects as heterotetramers; these were assigned PS3 at the moderate level. 17 variants had mild abnormalities with inconclusive predicted disease impacts and were not assigned any ACMG/AMP *in vitro* criteria. The electrophysiological results were largely consistent with the EHR phenotypes in the study (Table 1, Figure 4, Figure 5). For example, all 5 of the variants assigned PS3 at the strong level were present in at least 1 eMERGE-III carrier with an extreme or severe phenotype. The updated BS3 and PS3 criteria, as well as recently published hotspot criteria for the three studied genes^19^ were used to reclassify the 52 variants (Table 1, Table S13). 12 variants were reclassified: 3 VUS**→**LB, 7 VUS**→**LP, 1 VUS**→**P, and 1 LP**→**P. These data support the concept that genomic screening, EHR phenotypes, and follow-up *in vitro* functional studies can address individual ultra-rare variants, and assist with interpretation of VUS.

**Table 1:** Reclassification of VUS studied *in vitro*.

| Variant | # Carriers<br>(Affected/<br>Total) | Phenotype(s) | *In Vitro*<br>Result | Criteria<br>Added | ACMG<br>Class. |
| --- | --- | --- | --- | --- | --- |
| KCNH2 p.Ala244Val | 0/3 | — | Normal | BS3 | VUS→LB |
| KCNH2 p.Gly924Trp | 0/5 | — | Normal | BS3 PM1 | VUS |
| KCNH2 p.Pro1132Ala | 0/3 | — | Normal | BS3 PM1 | VUS |
| KCNH2 p.Pro1075Leu | 0/3 | — | Normal | BS3 PM1 | VUS |
| KCNH2 p.Glu682Asp | 1/1 | SCD | Normal | BS3 PM1 | VUS |
| KCNH2 p.Cys1117Ser | 1/1 | VT/VF, Syncope, AF,<br>Ad.AVB, SSS | Normal | BS3 PM1 | VUS |
| KCNQ1 p.Gly626Ser | 0/3 | — | Near Normal | BS3 PM1p | VUS→LB |
| KCNQ1 p.Arg181Leu | 0/1 | — | Normal | BS3 PM1 | VUS→LP |
| KCNQ1 p.Cys381Trp | 0/1 | — | Mild LOF | PM1 | VUS→LP |
| KCNQ1 p.Arg533Trp | 0/1 | — | Severe LOF | PS3m PM1 | VUS→LP |
| KCNQ1 p.Val280Glu | 1/1 | LQTS, Syncope, CIED | Severe LOF+DN | PS3 PM1 | VUS→P |
| KCNQ1 p.Gly186Asp | 1/1 | LQTS | Severe LOF+DN | PS3 PM1 | LP→P |
| KCNQ1 p.Ala31Val | 3/6 | Ad.AVB, LBBB, RBBB, PVCs;<br>SCD; PVCs, Syncope | Near normal | BS3 | VUS |
| KCNQ1 p.Gly57Ser | 3/9 | AF, Syncope; Syncope; AF,<br>CIED | Normal | BS3 | VUS |
| KCNQ1 p.Arg181His | 1/1 | PVCs | Near normal | BS3 PM1 | VUS |
| KCNQ1 p.Ser475Asn | 1/1 | Syncope | Near normal | BS3 PM1p | VUS |
| KCNQ1 p.Leu491Val | 1/1 | Syncope | Near normal | BS3 PM1p | VUS |
| KCNQ1 p.Thr495Pro | 1/1 | PVCs | Near normal | BS3 PM1p | VUS |
| KCNQ1 p.Glu449Lys | 1/3 | Syncope | Normal | BS3 PM1p | VUS |
| KCNQ1 p.Gly57Arg | 1/4 | AF | Near Normal | BS3 | VUS |
| KCNQ1 p.Asp388Asn | 0/1 | — | Normal | BS3 PM1 | VUS |
| KCNQ1 p.Glu290Lys | 2/4 | Syncope; AF | Mild LOF | PM1 | VUS |
| KCNQ1 p.Pro448Leu | 1/1 | Syncope | Mild LOF | PM1p | VUS |
| KCNQ1 p.Ser66Phe | 1/1 | AF, Syncope | Mild LOF |  | VUS |
| KCNQ1 p.Ala287Val | 1/1 | PVCs | GOF | PM1 | VUS |
| KCNQ1 p.Pro441Ser | 1/1 | AF | Mild LOF | PM1p | VUS |
| KCNQ1 p.Arg452Leu | 1/1 | PVCs | Mild LOF | PM1p | VUS |
| KCNQ1 p.Gln531Glu | 1/1 | Syncope | Mild LOF | PM1 | VUS |
| KCNQ1 p.Lys398Arg | 1/2 | LBBB | Mild LOF | PM1p | VUS |
| KCNQ1 p.Asp488Glu | 1/2 | AF, Ad. AVB, LBBB, SSS,<br>VT/VF | Mild LOF | PM1p | VUS |
| KCNQ1 p.Gly635Arg | 1/3 | AF | Mild LOF | PM1p | VUS |
| KCNQ1 p.Gly57Val | 1/4 | AF, CIED | Mild LOF |  | VUS |
| KCNQ1 p.Pro67Arg | 0/2 | — | GOF |  | VUS |
| KCNQ1 p.Thr444Met | 0/2 | — | Mild LOF | PM1p | VUS |
| KCNQ1 p.Ile574Val | 0/3 | — | Mild GOF | PM1 | VUS |
| KCNQ1 p.Arg519His | 2/4 | Syncope; Syncope | Severe LOF | PS3m PM1 | VUS |
| KCNQ1 p.Asp649Asn | 0/2 | — | Mild LOF | PM1p | LB |
| SCN5A p.Pro1123Leu | 0/3 | — | Normal | BS3 | VUS→LB |
| SCN5A p.Phe1794Ile | 1/1 | AF, Ad.AVB, SSS, PVCs,<br>Syncope, CIED | Partial LOF | PS3 PM1p | VUS→LP |
| SCN5A p.Gly1605Asp | 1/1 | AF, VT/VF | GOF | PS3 PM1 | VUS→LP |
| SCN5A p.Asp1741Asn | 1/1 | AF, VT/VF | GOF | PS3 PM1 | VUS→LP |
| SCN5A p.Ile848Phe | 1/3 | SSS, AF, PVCs | Mild LOF | PM1 | VUS→LP |
| SCN5A p.Ala662Ser | 0/4 | — | Normal | BS3 | VUS |
| SCN5A p.Gly552Trp | 0/4 | — | Normal | BS3 | VUS |
| SCN5A p.Lys1500Asn | 1/1 | AF, Ad.AVB, VT/VF, PVCs,<br>Syncope, CIED* | Normal | BS3 | VUS |
| SCN5A p.Ser835Ala | 1/1 | AF | Normal | BS3 PM1 | VUS |
| SCN5A p.Tyr1228His | 1/1 | Syncope | Normal | BS3 PM1 | VUS |
| SCN5A p.Ser172Thr | 1/1 | AF, PVCs, LQTS, Syncope | Normal | BS3 PM1 | VUS |
| SCN5A p.Gly1084Ser | 1/1 | AF, PVCs, LQTS, Syncope | Normal | BS3 | VUS |
| SCN5A p.Asn291His | 1/2 | AF, LQTS, Syncope | Normal | BS3 PM1 | VUS |
| SCN5A p.Ile94Val | 3/3 | AF; AF, PVCs; LQTS, PVCs | Normal | BS3 PM1 | VUS |
| SCN5A p.Gln245Lys | 1/1 | AF, SSS, VT/VF, PVCs,<br>Syncope | Normal | BS3 PM1p | VUS |

\*Clinical genetic testing indicated that this participant also has a pathogenic variant in *DSP* p.Arg1269Ter and a VUS in *JUP* p.Ser323Gly. Abbreviations: AF—atrial fibrillation, Ad.AVB—advanced atrioventricular block, CIED—cardiovascular implantable electronic device, LOF—loss of function, GOF—gain of function, DN—dominant negative, LQTS—long QT, PVCs—premature ventricular contraction, SCD: sudden cardiac death, SSS—sick sinus syndrome, VT/VF—ventricular tachycardia/ventricular fibrillation, VUS—variant of uncertain significance, P—pathogenic, LP—likely pathogenic, LB—likely benign. The criteria added column indicates new ACMG criteria that were applied based on *in vitro* functional and hotspot data. The PS3 criterion was applied at the strong (PS3) or moderate (PS3m) level. The PM1 criterion was applied at the moderate (PM1) or supporting (PM1p) level. The full set of applied ACMG criteria and additional variant data, is presented in Table S13.

## Discussion

### Association between P/LP and arrhythmia phenotypes

Using a large unselected cohort, we found associations between P/LP variants and arrhythmia phenotypes. These associations were strongest with LQTS, especially the potassium channel genes *KCNQ1, KCNH2*, and *KCNE1*, with adjusted odds ratios between 20-25. Variants in the potassium channel genes *KCNQ1* and *KCNH2* are the commonest cause of LQTS, responsible for approximately 80% of cases.^21^ *KCNE1* encodes an auxiliary subunit of an ion channel responsible for the repolarizing current I_Ks_, and was recently designated by the ClinGen consortium as only having “limited evidence” for LQTS association.^21^ Our results suggest that although *KCNE1* P/LP variants may be rare, in part due to the small size of the gene, at least some *KCNE1* variants are strongly associated with LQTS. *SCN5A* and *LMNA* P/LP variants had pleiotropic effects: *SCN5A* variants were associated with syncope and premature ventricular contractions, and *LMNA* variants were associated with 7 arrhythmia phenotypes. Using ECG intervals as a quantitative readout of variant effect, we also observed associations between P/LP variants in *KCNH2* and *KCNQ1* and QTc prolongation and variants in *SCN5A* with slower cardiac conduction (PR and QRS prolongation), consistent with these channels’ effects on the cardiac conduction system.

Previous work from the eMERGE consortium failed to detect a high rate of arrhythmia diagnoses in carriers of incidental variants in *SCN5A* or *KCNH2*.^22^ Three main reasons may explain the difference between the previous and current studies. First, the current dataset is much larger (21,846 vs 2,022) and includes additional genes (10 genes vs. 2), improving the power to detect variant associations. Second, the current study included return of results to the participants and follow-up examination and diagnoses. Third, since the widespread adoption of the ACMG/AMP framework,^18^ variant annotation practices appear to have improved in consistency, and many variants have been reclassified.^23^

### Incomplete penetrance

Incomplete penetrance—the phenomenon that only a fraction of P/LP variant carriers have a disease-related phenotype—has been well-described for many arrhythmia variants studied in large families. For example, large family studies showed a penetrance of 25%-41% for LQTS,^24,25^ 47%-65% for BrS,^26,27^ and 76%-92% for *LMNA*-related cardiomyopathy and arrhythmia.^11,28^ In our study, of the 53 participants with returned P/LP variants who were invited for detailed follow-up examination, 38% had arrhythmia diagnoses and 55% had documented arrhythmia phenotypes. Therefore, it appears that we can apply the lessons of the family studies to the general population, although the penetrance in unselected populations may be lower. Several variables help explain incomplete penetrance for arrhythmia disorders, such as genetic background (including common polygenic risk), sex, and age; environmental influences such as drug exposures and electrolyte derangements can also elicit a phenotype in susceptible individuals.^29-31^ Future precision medicine efforts could integrate these variables with P/LP variant status to better identify individuals at high risk of developing serious arrhythmias.

### In vitro functional evaluation and variant reclassification

Ultra-rare VUS are plausible disease-causing variants and were detected in a large fraction (8.4%) of participants. VUS were detected at a higher rate in self-reported non-White participants compared to White participants, similar to previous findings for hereditary cancer-predisposing genes.^32^ However, only *LMNA* VUS had any significant arrhythmia associations, with extreme and severe phenotypes. 116 ultra-rare VUS detected in at least 1 individual with a severe or extreme arrhythmia phenotype, but the EHR code-based data alone were not sufficient to reclassify these VUS because these variants typically were present in only one or a few participants. To further decipher these variants’ disease risk, we used automated patch clamp recording, an emerging technology that enables the high-throughput electrophysiological study of ion channel variants.^6,9,33^ Based on *in vitro* functional testing and recently published hotspot criteria,^19^ we were able to reclassify 12 of 52 examined variants. Future analyses studying even larger cohorts, independently or combined with high-throughput *in vitro* characterization^6,34^ promise to further discern the disease risk of individual ultra-rare variants.

### Prospects for arrhythmia genomic screening

Out of the 21,846 participants in this study, P/LP variants in arrhythmia genes were returned to 53 participants. These individuals had a high rate of arrhythmia phenotypes (55%) and inherited arrhythmia diagnoses (38%), a majority of which were made following return of variant results. These data and the broader EHR-derived arrhythmia associations discussed above indicate that incidental P/LP variants in selected arrhythmia genes are associated with increased risk of arrhythmias. Population genomic screening has the potential to positively impact public health, and has begun to be deployed for some diseases such as hereditary cancer syndromes and familial hypercholesterolemia.^35^ Our results suggest that arrhythmia genomic screening may also be merited, especially in the context of simultaneous screening for multiple Mendelian disease genes. However, future work is needed to better understand the accuracy and cost-effectiveness of arrhythmia genomic screening.

### Limitations

This study used EHR phenotypes for some analyses, which risks misdiagnosing or underdiagnosing certain clinical phenotypes. This applies in particular to specific arrhythmia phenotypes, such as BrS or CPVT, which may be challenging to ascertain from the EHR and may require special diagnostic testing and interpretation. Despite consenting for the genomic screening study, 21/51 participants who were returned the result of a P/LP variant did not seek follow-up care with a cardiology specialist or genetic counselor. Therefore, penetrance of the P/LP variants in this study might be underestimated due to incomplete ascertainment. P/LP variants in three genes (*KCNE1, KCNE2*, and *KCNJ2*) and specific variants in other genes were present in a low number of participants, limiting the power of analyses using these genes or variants. Estimates of penetrance might also be improved with family history information, which was not available in eMERGE-III.

## Conclusions

In a cohort of more than 20,000 unselected participants, P/LP variants in arrhythmia genes were associated with arrhythmia phenotypes. After return of results of P/LP variants, inherited arrhythmia diagnoses were made in nearly half of carriers. Follow-up *in vitro* functional evaluation enabled VUS reclassification. In large cohorts, rare variants in arrhythmia genes can be characterized by integrating genomic screening, EHR phenotypes, and *in vitro* functional studies.

## Methods

### Study participants

We studied the prospective Electronic Medical Records and Genomics (eMERGE-III) cohort consisting of 21,846 individuals recruited from 10 academic medical centers in the United States.^16,17^ There were 24,956 individuals recruited for the eMERGE-III study but we excluded 410 from the analyses presented here due to incomplete age and/or sex information, 2,498 (all from a single site) because they were enrolled based on the results of previous sequencing, and 202 because their indications for enrollment were arrhythmia, cardiomyopathy, or heart failure. All participants consented to have P/LP sequencing results reported to them and to their EHR. This project was approved by the Institutional Review Boards of the individual centers. Individuals were enrolled between January 2016 and January 2018.

### Sequencing and variant annotation

Individuals were sequenced for the complete coding regions of 109 genes, including the 10 arrhythmia-associated genes reported here, using a custom Illumina capture platform. A detailed description of this sequencing project and variant calling pipeline has been previously published.^16^ P/LP variants in the five arrhythmia genes that were returned (*KCNE1, KCNH2, KCNQ1, LMNA*, and *SCN5A*) were annotated by the sequencing centers using ACMG/AMP criteria.^18^ Variants in the other five genes (*ANK2, CACNA1C, KCNE2, KCNJ2*, and *RYR2*) were evaluated using ClinVar annotations^36^ and manual review using ACMG/AMP criteria.^18^ No gain of function *RYR2* P/LP variants associated with CPVT risk were detected or returned, consistent with the low prevalence of CPVT.^12^ However, loss of function *RYR2* variants, recently associated with sudden arrhythmic death,^37^ were also classified as P/LP and included in the EHR-based analyses. Exome and genome sequencing counts from gnomAD v.2.1.1 were used to calculate a merged gnomAD allele frequency.^38^ Principal components of ancestry were determined from an analysis of ancestry informative markers included on the eMERGE-III platform as previously described.^39^

### Return of results

A description of the overall return of results (RoR) for the eMERGE-III project has been published.^17^ Briefly, each site decided which P/LP variants to return, largely following the ACMG list of recommended genes^40^ (*KCNH2, KCNQ1, SCN5A*, and *LMNA*), but also including *KCNE1* variants for some sites. As described above, no *RYR2* P/LP CPVT variants were identified so none were returned. VUS were not returned. Two variants, *KCNE1* p.Asp85Asn (rs1805128) and *KCNE2* p.Met54Thr (rs74315447) were classified as “risk factor” variants.^21^ These variants were returned by some sites, but they were not included in the analyses presented here. Return of results occurred between July 2017 and May 2019, and there was 1 year of follow-up for these participants. During this follow-up period, participants with P/LP variants in arrhythmia genes were contacted and encouraged to pursue a genetic counseling/cardiology follow-up visit. To capture the impact of return of sequencing results, three RedCap surveys were filled out by specialist reviewers: immediately following return of results, after 6 months, and after 1 year. These surveys consisted of manual chart review to ascertain which patient phenotypes and diagnoses were available at each of the three time periods. Inherited arrhythmia diagnoses were adjudicated by clinicians at each site, either during a specialist follow-up visit or by diagnostic codes in the EHR.

### Comparison by variant status

Participants heterozygous for P/LP variants were defined as described above. Participants heterozygous for VUS were defined as individuals who 1) did not have a P/LP variant in the 10 sequenced arrhythmia genes and 2) were a heterozygous for a VUS with a minor allele frequency <2.5e-5 in the gnomAD database, following a previously suggested cutoff for Mendelian arrhythmia variants.^20^ Non-carriers were defined as individuals not in the first two groups for any of the 10 arrhythmia-associated genes. All demographic comparisons between these groups were made with two-sided Fisher’s Exact Tests, implemented in R with the function *fisher*.*test* with the exception of age, which was analyzed as a continuous variable using a Kruskal-Wallis test using the R function *kruskal*.*test*.

### Generation of phenotypes from EHR codes

Arrhythmia phenotypes were extracted from the core phenotype set of eMERGE-III, using International Classification of Diseases codes (ICD9, ICD10) and Current Procedural Terminology (CPT) codes from EHRs. The 11 arrhythmia-related phenotypes were long QT syndrome (LQTS), sudden cardiac death (SCD) [also including sudden unexplained death and sudden infant death syndrome], ventricular tachycardia/ventricular fibrillation (VT/VF), advanced atrioventricular block (Ad. AVB) [including second degree AVB, bifascicular or trifascicular AVB, high-grade or complete heart block], presence of cardiovascular implantable electronic devices (CIEDs), sick sinus syndrome (SSS), atrial fibrillation/atrial flutter (AF/AFL), left bundle branch block (LBBB), premature ventricular contractions (PVCs), right bundle branch block (RBBB), and syncope (unspecified). A full description of the associated codes that were used to generate these code-based phenotypes is presented in File S1.

Arrhythmia phenotypes were grouped by severity as follows:

1. Extreme: LQTS, SCD, VT/VF.
2. Severe: SSS, CIEDs, Ad. AVB.
3. Moderate: AF/AFL.
4. Mild: PVCs, LBBB, RBBB, or syncope.

### Electrocardiographic parameters

Electrocardiographic (ECG) data (PR, QRS, QT, heart rate) were available from the EHR from 4 sites for all individuals (P/LP carriers, VUS carriers and non-carriers). Bazett’s formula was used to derive the corrected QT interval (QTc) from the QT and heart rate (QTc=QT/(RR^0.5), QT measured in milliseconds and RR in seconds). ECGs without a reported heart rate, QT, or QRS were excluded. ECGs with outlier intervals (QT ≤ 150 or >1000 ms, QRS ≤ 20 ms or >1000 ms, or PR ≤ 40 ms or >1000ms) were also excluded. To avoid confounding effects on these intervals from tachycardia and bradycardia, only ECGs with heart rate between 50-100 beats per minute were included in the analysis. If a participant had more than one ECG meeting inclusion criteria, median values for each interval were used in subsequent analyses. For individuals with returned variants, case review surveys included data derived from the last available ECG. For the maximum QTc column in Table S11, if individuals had ECG data available from both case review surveys and from the EHR, the maximum QTc from either source was used.

### Phenotype enrichment analyses

Associations between carrier status and the 11 EHR code-derived phenotypes were examined by comparing 1) P/LP heterozygotes to non-carriers and 2) VUS heterozygotes to non-carriers, using the criteria defined above. These associations were evaluated using two methods. The first method was logistic regression, implemented in Stata. The model predicted arrhythmia phenotype from genetic case or control status, and included age, sex, study site, and first 10 principal components of ancestry as covariates. Phenotype and genetic case or control status were binary variables. The p-values and effect sizes were determined using the Stata function *logistic*. The second method used the Optimized Sequence Kernel Association Test (SKAT-O),^41^ implemented using *rvtests* software,^42^ using the same covariates as above. For Figure 1, the associations between variant status and “severe” or “extreme” phenotypes were also examined by logistic regression with similar covariates as above comparing 1) P/LP heterozygotes to non-carriers and 2) VUS heterozygotes to non-carriers.

Associations between variant status and ECG intervals were determined by linear regression implemented in Stata. The model predicted ECG intervals as continuous variables from genetic case or control status, and included age, sex, site, and 10 principal components of ancestry as covariates.

For all these sets of tests, false discovery rates (FDRs) were calculated from the nominal p-values using the R function *p*.*adjust* using the “Benjamini & Hochberg” option. FDRs less than 0.1 were considered statistically significant.

### EHR analyses of individual variants

Variants were pooled according to multiple criteria in separate analyses (e.g. by gene, by clinical annotation, by individual variant). For individual variant analyses, ultra-rare (gnomAD allele frequency <2.5e-5)^20^ variants with at least three carriers in the cohort or at least one carrier with a severe or extreme arrhythmia phenotype (as defined above) were analyzed. The frequencies of phenotypes in these subgroups were calculated using the four arrhythmia phenotype groups described above.

### Electrophysiological evaluation of variants

A subset of variants in *KCNH2, KCNQ1*, and *SCN5A* were selected for *in vitro* study using the SyncroPatch 384PE or 768PE automated patch clamp devices. *KCNH2* and *SCN5A* variants were studied as previously described.^6^ Briefly, *KCNH2* and *SCN5A* variants were generated with QuikChange Multi Kit (Agilent) in a “zone” plasmid (fragment of full length gene) and subcloned into a *SCN5A*:IRES:mCherry-blasticidinR or *KCNH2*:IRES:mCherry-blasticidinR plasmid. All mutagenesis primers are listed in Table S14. *KCNH2* and *SCN5A* mutant or wildtype plasmids were integrated into a HEK293 landing pad negative selection line,^43^ selected with AP1903 (MedChem Express) and blasticidin (ThermoFisher), validated by flow cytometry, and patch clamped on the SyncroPatch 384PE as previously described.^6^ For *SCN5A*, variants with normalized peak current density <10%, 10-50%, 50-75%, 75-125%, >125% were considered loss of function, partial loss of function, mild loss of function, normal, and gain-of-function, respectively. Late current measurements for p.Gly1605Asp were confirmed by manual patch-clamp, as previously described^44^ using the cell lines described above. Cutoffs for *KCNH2* were identical as the *SCN5A* cutoffs above, except KCNH2 tail current was used. *KCNQ1* variants were generated and studied in combination with KCNE1 in either the homotetrameric or heterotetrameric state (coexpressed 1:1 with WT KCNQ1) in Chinese Hamster Ovary cells on the SyncroPatch 768PE as previously described.^9^ For *KCNQ1*, variants with normalized peak current density <25%, 25-75%, 75-90%, 90-110%, 110-125%, 125-150%, >150% compared with WT channels were considered loss of function, partial loss of function, near-normal, normal, near-normal, mild gain-of-function, and severe gain-of-function, respectively. For the three genes, additional parameters were collected and examined, *i*.*e*. voltage dependence of activation/inactivation, activation/deactivation speeds, and recovery from inactivation, but none of the variants had large enough changes in these parameters compared to wildtype to modify the peak/tail current derived cutoffs mentioned above. The full dataset of measured patch clamp parameters for each variant is presented in File S2.

Following functional evaluation, variants were reclassified using ACMG/AMP classification criteria. A full description of the ACMG/AMP classification criteria used is detailed in the Supplemental Methods.

## Data Availability

The eMERGE-III datasets are publicly available in the dbGaP repository under phs001616.v2.p2 and access can also be requested on the eMERGE website https://emerge-network.org.

## Author Contributions

AMG, GD, CMS, CGV, RRD, EHF, OD, NS, JAP, TY, AM, QSW, LLR, ORK, SB, ZTY, DWM, BMK, IJK, GPJ, ASG, EBL, TM, JZL, DS, BN, TA, RS, AS, CL, CW, GH, JDR, EMM, WKC, DSC, KAL, HH, PS, SS, JG, the eMERGE Network, JD, WQW, ALG, BMS, and DMR participated in dataset acquisition. AMG, GD, CMS, CGV, RRD, TY, JDM, ALG, MBS, DMR analyzed the data. AMG, GD, JDM, SLVD, YW, GPJ, TAM, WKC, STM, ALG, MBS, and DMR wrote the manuscript. All authors reviewed and approved the final manuscript.

## Acknowledgements

This project used datasets obtained for the eMERGE Network (Phase III). This phase of the eMERGE Network was initiated and funded by the NHGRI through the following grants: U01HG8657 (Kaiser Permanente Washington/University of Washington); U01HG8685 (Brigham and Women’s Hospital); U01HG8672 (Vanderbilt University Medical Center); U01HG8666 (Cincinnati Children’s Hospital Medical Center); U01HG6379 (Mayo Clinic); U01HG8679 (Geisinger Clinic); U01HG8680 (Columbia University Irving Medical Center); U01HG8684 (Children’s Hospital of Philadelphia); U01HG8673 (Northwestern University); U01HG8701 (Vanderbilt University Medical Center serving as the Coordinating Center); U01HG8676 (Partners Healthcare/Broad Institute); and U01HG8664 (Baylor College of Medicine). This research was also funded by NIH grants K99 HG010904 (AMG), R01 HL149826 (DMR), NIH R01 HL128075 and an AHA SFRN (EMM), R01 HL122010 (ALG).

Electrocardiographic data at Vanderbilt University Medical Center were obtained using Vanderbilt’s Synthetic Derivative. The Synthetic Derivative resource is supported by CTSA award No. UL1TR000445 from the National Center for Advancing Translational Sciences. The contents of this publication are solely the responsibility of the authors and do not necessarily represent official views of the National Center for Advancing Translational Sciences or the National Institutes of Health.

