## Supplement for "Arrhythmia variant associations and reclassifications in the eMERGE-III sequencing study"

**in the eMERGE-III sequencing study**

**Supplement**

<u>**Table of Contents**</u>


**Supplemental Methods**

**Table S1:** Participant characteristics
**Table S2:** Genes sequenced in this study
**Table S3:** Associations between P/LP heterozygotes and EHR code-derived phenotypes
**Table S4:** Associations between VUS heterozygotes and EHR code-derived phenotypes
**Table S5:** SKAT-O burden test analysis (P/LP variants)
**Table S6:** SKAT-O burden test analysis (VUS)
**Table S7:** Pre-2017 associations between P/LP heterozygotes and EHR code-derived phenotypes
**Table S8:** Associations between P/LP heterozygotes and EHR code-derived phenotypes in self-reported White participants
**Table S9:** Differences in ECG intervals obtained from the EHR as a function of P/LP status
**Table S10:** Differences in ECG intervals obtained from the EHR as a function of VUS carrier status
**Table S11:** Participants with P/LP variants diagnosed with inherited arrhythmia syndromes
**Table S12:** Changes in clinical management in participants with returned P/LP variants
**Table S13:** Additional properties of variants studied *in vitro*
**Table S14:** Mutagenesis primers

**Figure S1:** Penetrance of variant carriers by variant (*ANK2*, *CACNA1C*, *KCNE2*, *KCNJ2*, and *RYR2*)
**Figure S2:** *In vitro* functional characterization of *KCNH2* variants

**File S1:** Diagnostic and procedure codes used to generate EHR phenotypes (.docx)
**File S2:** All measured patch clamp parameters for each variant. (.xlsx)

**Literature Cited**

**Supplemental Methods**

### ACMG-AMP Classification

Variants were classified according to ACMG-AMP classification criteria.[1] The following criteria were implemented:

**BS3 and PS3:** BS3 (well-established functional studies show no deleterious effect) was applied if the variants had normal or near-normal *in vitro* function as defined in the Methods. For *KCNQ1*, BS3 was applied only if the variants had normal or near-normal *in vitro* function in both homotetrameric and heterotetrameric experiments. PS3 (well-established functional studies show a deleterious effect) was applied at the strong level for *SCN5A* variants that had GOF, partial LOF, or LOF. PS3 was applied at the strong level for *KCNQ1* variants that had severe LOF and a strong dominant negative effect in the heterotetrameric experiment. PS3 was applied at the moderate level for *KCNQ1* variants that had severe LOF in the homotetrameric experiment but did not have a strong dominant negative effect in the heterotetrameric experiment. All other cases were not assigned either BS3 or PS3.

**BP4 and PP3:** BP4 (multiple lines of computational evidence support no impact) and PP3 (multiple lines of computational evidence support a deleterious effect) were implemented using the online tool CardioClassifier.[2] CardioClassifier determined computational predictions with 8 algorithms (SIFT, MutationTaster, MutationAssessor, Polyphen-VAR, LRT, FATHMM, Grantham, CADD). It then determined if all predictions or all except one met a consensus that the variant was predicted to be deleterious (PP3) or non-deleterious (BP4). If there was not such a consensus, BP4 and PP3 were not applied.

**PS4:** PS4 was implemented for *KCNQ1*, *KCNH2*, and *SCN5A* using recently published[3] criteria for these genes.

**PP5 and BP6:** PP5 (reputable source reports pathogenic) and BP6 (reputable source reports benign) were implemented using ClinVar.[4] Only reports from commercial genetic testing companies after 2016 were included.

**PM1:** PM1 (mutational hotspot or well-studied functional domain) was implemented for *KCNQ1*, *KCNH2*, and *SCN5A* following a recently published set of recommended hotspot boundaries[3] for those 3 genes. However, this criterion was only implemented at the moderate or supporting level; variants that were recommended in the previous study to be PM1 (strong) were implemented here at the PM1 (moderate) level.

**BA1, BS1, and PM2:** BA1/BS1 (Minor allele frequency too high for the relevant disorder) and PM2 (Absent from control cohort with >1,000 individuals) were assigned based on gnomAD minor allele frequencies.[5] Following a previous study,[3] variants with a minor allele frequency below 1e-5 in gnomAD were assigned PM2, variants with MAFs between 1e-5 and 5e-5 were not assigned any of these criteria, variants with MAFs between 5e-5 and 0.05 were assigned BS1, and variants with MAF>0.05 were assigned BA1.

**PP2:** All missense variants in this study were assigned PP2 (missense variant in gene for which missense variant is a common mechanism of disease).

**PVS1:** PVS1 (Predicted null variant if loss of function is a disease mechanism) were applied to frameshift, nonsense, and canonical splice site (1-2 bp from exon) variants. Variants of these classes near the N-terminal end of the protein, however, especially variants in the last exon or the last 50 bp of the penultimate exon were not considered to meet PVS1, unless evidence was available that these were null variants.

**PM5:** PM5 (novel missense variant at amino acid position with different pathogenic missense variant) was implemented with ClinVar, as in PP5 and BP6 (above).

**PS4:** PS4 (prevalence in cases statistically enriched over controls) for *KCNQ1*, *KCNH2*, and *SCN5A* was implemented using a recently published dataset and analysis of LQTS and BrS case and control variant prevalence.[3]

**Table S1: Participant characteristics**

| Characteristic | Overall Cohort (n= 21,846) | Non-Carrier (n=19,885) | P/LP Carrier (n=123) | VUS Carrier (n=1,838) | *P* Value |
|---|---|---|---|---|---|
| Age, median (IQR), years | 58 (26-70) | 58 (26-70) | 53 (24-66) | 55 (24-68) | 5.0e-5^ |
| Age < 18y, no. (%) | 2,377 (10.9%) | 2,134 (10.7%) | 18 (14.6%) | 225 (12.2%) | 0.053 |
| Female sex -no. (%) | 11,399 (52.2%) | 10,372 (52.2%) | 65 (52.8%) | 962 (52.3%) | 0.979 |
| **Self-reported race - no (%)** | | | | | |
| White | 15,238 (69.8%) | 14,032 (70.6%) | 80 (65.0%) | 1,126 (61.3%) | 1.8e-15 |
| Black | 3,849 (17.6%) | 3,426 (17.2%) | 30 (24.4%) | 393 (21.4%) | 9.7e-6 |
| Asian | 1,556 (7.1%) | 1,348 (6.8%) | 6 (4.9%) | 202 (11.0%) | 1.1e-9 |
| Other* | 1,203 (5.5%) | 1,079 (5.4%) | 7 (5.7%) | 117 (6.4%) | 0.228 |
| **Ethnicity - no (%)** | | | | | |
| Non-Hispanic | 20,071 (91.9%) | 18,266 (91.9%) | 119 (96.7%) | 1686 (91.7%) | 0.120 |
| Hispanic | 1,441 (6.6%) | 1,316 (6.6%) | 2 (1.6%) | 123 (6.7%) | 0.057 |
| Unknown | 334 (1.5%) | 303 (1.5%) | 2 (1.6%) | 29 (1.6%) | 0.868 |

^*P* value was calculated from a Kruskal-Wallis test. All other *P* values were calculated from Fisher's Exact tests comparing non carrier, P/LP carrier, and VUS carrier groups. *Other race includes Native Hawaiian, American Indian, and no reported race. P/LP—Pathogenic/likely pathogenic; VUS—Variant of Uncertain Significance.

**Table S2: Genes sequenced in this study**

| Gene | Description | Disease |
|---|---|---|
| *ANK2* | Ankyrin 2 | LQTS Type 4 |
| *CACNA1C* | $Ca^{2+}$ channel | LQTS Type 8 |
| *KCNE1* | $K^{+}$ channel | LQTS Type 5 |
| *KCNE2* | $K^{+}$channel | LQTS Type 6 |
| *KCNH2* | $K^{+}$channel | LQTS Type 2 |
| *KCNJ2* | $K^{+}$channel | LQTS Type 7 |
| *KCNQ1* | $K^{+}$channel | LQTS Type 1 |
| *LMNA* | Lamin A/C | Cardiomyopathy, conduction disease, progeria |
| *RYR2* | $Ca^{2+}$ channel (ryanodine release) | CPVT Type 1 |
| *SCN5A* | $Na^{+}$ channel | LQTS Type 3, BrS Type 1 |

LQTS = Long QT Syndrome, CPVT = Catecholaminergic Polymorphic Ventricular Tachycardia, BrS = Brugada Syndrome

**Table S3: Associations between P/LP heterozygotes and EHR code-derived phenotypes**

| Phenotype | All genes (n=123) | *ANK2* (n=16) | *KCNE1* (n=9) | *KCNH2* (n=7) | *KCNQ1* (n=30) | *LMNA* (n=7) | *RYR2* (n=31) | *SCN5A* (n=25) |
|---|---|---|---|---|---|---|---|---|
| **Extreme** | | | | | | | | |
| LQTS | **11.7 (1.4e-13)*** | 0 (1) | **24.9 (1.5e-4)*** | **20.1 (0.001)*** | **24.4 (3.6e-11)*** | 0 (1) | 0 (1) | 4.6 (0.15) |
| SCD | 0 (1) | 0 (1) | 0 (1) | 0 (1) | 0 (1) | 0 (1) | 0 (1) | 0 (1) |
| VT/VF | **2.7 (0.025)*** | 5.2 (0.15) | 0 (1) | 0 (1) | 0 (1) | **25.6 (0.001)*** | 4.9 (0.050) | 4.6 (0.053) |
| **Severe** | | | | | | | | |
| Advanced AV block | 2.1 (0.16) | 6.3 (0.13) | 0 (1) | 0 (1) | 0 (1) | **13.6 (0.026)*** | 4.9 (0.046) | 2.7 (0.34) |
| CIEDs | 1.7 (0.38) | 0 (1) | 0 (1) | 0 (1) | 0 (1) | **18.1 (0.016)*** | 5.4 (0.038) | 0 (1) |
| Sick Sinus | **3.1 (0.013)*** | 0 (1) | 0 (1) | 0 (1) | 2 (0.52) | **36.8 (1.0e-4)*** | 4.8 (0.06) | 2.2 (0.45) |
| **Moderate** | | | | | | | | |
| AF/AFL | 1.0 (0.91) | 0 (1) | 0.8 (0.82) | 0 (1) | 0.3 (0.24) | **22.9 (0.003)*** | 1.6 (0.48) | 0.9 (0.87) |
| **Mild** | | | | | | | | |
| LBBB | 1.8 (0.25) | 0 (1) | 0 (1) | 0 (1) | 0 (1) | **35.1 (4.6e-4)*** | 2.3 (0.44) | 2.3 (0.42) |
| PVCs | 1.5 (0.31) | 2.4 (0.45) | 0 (1) | 0 (1) | 0 (1) | 5.5 (0.143) | 3.5 (0.036) | **4.3 (0.016)*** |
| RBBB | 1.4 (0.53) | 5.0 (0.16) | 0 (1) | 0 (1) | 1.2 (0.88) | **10.3 (0.044)** | 1.4 (0.78) | 1.8 (0.57) |
| Syncope | 1.8 (0.035) | 2.1 (0.39) | 2.2 (0.35) | 1.2 (0.86) | 0.3 (0.23) | **12.3 (0.003)*** | 0.8 (0.76) | **6.0 (1.4e-4)*** |

Logistic regression adjusted for sex, 10 principal components of ancestry, age, and site. Entries indicate Odds Ratio (p-value). *CACNA1C*, *KCNE2* and *KCNJ2* had 4, 1, and 0 individuals with pathogenic/likely pathogenic variants, respectively, all without any arrhythmia codes, so odds ratios and p-values were not calculated. Bold: Nominal p-value <0.05, unadjusted for multiple testing. * indicates false discovery rate < 0.1 (adjusted for p-values in this table). Abbreviations: LQTS— long QT syndrome, SCD— sudden cardiac death, VT/VF— ventricular tachycardia/ventricular fibrillation, CIED—cardiovascular implantable electronic devices, AF/AFL—atrial fibrillation/atrial flutter, LBBB—left bundle branch block, PVC—premature ventricular contraction, RBBB—right bundle branch block.

**Table S4. Associations between VUS heterozygotes and EHR code-derived phenotypes**

| Phenotype | All genes (n=1838) | *ANK2* (n=493) | *CACNA1C* (n=219) | *KCNE1* (n=23) | *KCNH2* (n=177) | *KCNJ2* (n=27) | *KCNQ1* (n=109) | *LMNA* (n=80) | *RYR2* (n=528) | *SCN5A* (n=284) |
|---|---|---|---|---|---|---|---|---|---|---|
| **Extreme** | | | | | | | | | | |
| LQTS | 0.8 (0.369) | 0.9 (0.909) | 0.4 (0.382) | 0 (1) | 0 (1) | 0 (1) | 1.8 (0.411) | **3.7 (0.032)** | 0.5 (0.28) | 1.3 (0.63) |
| SCD | 1.2 (0.562) | 1.4 (0.476) | 0.6 (0.643) | 0 (1) | 1.5 (0.583) | 0 (1) | 1.4 (0.746) | 0 (1) | 1.2 (0.636) | 1 (0.952) |
| VT/VF | 1 (0.814) | 0.8 (0.531) | 0.9 (0.849) | 0 (1) | 0.8 (0.706) | 0 (1) | 0.4 (0.41) | **3.4 (0.012)** | 1.1 (0.746) | 1 (0.934) |
| **Severe** | | | | | | | | | | |
| Advanced AV block | **0.6 (0.046)** | 0.4 (0.083) | 0.3 (0.171) | 0 (1) | 0.7 (0.555) | 3 (0.295) | 1 (0.958) | **3.8 (0.007)** | 0.5 (0.181) | 0.4 (0.204) |
| CIEDs | 0.8 (0.366) | **0.1 (0.028)** | 0.9 (0.854) | 0 (1) | 1.1 (0.869) | 3.5 (0.237) | 1.6 (0.408) | **4.3 (0.003)** | 0.6 (0.219) | 1 (0.972) |
| Sick Sinus | 0.9 (0.559) | 1 (0.88) | 1.2 (0.732) | 0 (1) | 0.7 (0.566) | 2.1 (0.479) | 0.4 (0.437) | 1.2 (0.821) | 0.5 (0.118) | 1.2 (0.599) |
| **Moderate** | | | | | | | | | | |
| AF/AFL | 1.1 (0.291) | 1 (0.906) | 0.7 (0.264) | 1.1 (0.89) | 0.9 (0.817) | 0.4 (0.39) | 1 (0.949) | **2.3 (0.014)** | 1.2 (0.293) | 1.1 (0.719) |
| **Mild** | | | | | | | | | | |
| LBBB | 0.9 (0.51) | 0.9 (0.837) | 0.9 (0.878) | 0 (1) | 0 (1) | 0 (1) | 1.3 (0.62) | 2.2 (0.139) | 0.9 (0.64) | 0.7 (0.561) |
| PVCs | 1.1 (0.596) | 1.1 (0.517) | 1.3 (0.388) | 0.9 (0.947) | 0.9 (0.834) | 0 (1) | 1 (0.975) | 1.8 (0.163) | 1 (0.846) | 1 (0.892) |
| RBBB | **0.7 (0.019)** | 0.6 (0.146) | 0.7 (0.473) | 0 (1) | 0.4 (0.258) | 0 (1) | 0.3 (0.259) | 2.5 (0.061) | 0.7 (0.301) | 0.5 (0.236) |
| Syncope | 1 .0 (0.915) | 1.1 (0.667) | 0.9 (0.732) | 2.3 (0.151) | 0.9 (0.839) | 0.9 (0.922) | **1.8 (0.043)** | 0.4 (0.117) | 1 (0.821) | 0.8 (0.289) |

Logistic regression (ultra-rare VUS heterozygotes vs. non-carrier) adjusted for sex, 10 principal components of ancestry, age, and site. Values indicate Odds Ratio (p value). There were only 3 *KCNE2* VUS heterozygotes so this gene was excluded from the table. Bold: Nominal p-value <0.05, unadjusted for multiple testing. None of these associations had a false discovery rate < 0.1 (adjusted for all p-values in this table). Abbreviations: LQTS— long QT syndrome, SCD— sudden cardiac death, VT/VF— ventricular tachycardia/ventricular fibrillation, CIED— cardiovascular implantable electronic devices, AF/AFL—atrial fibrillation/atrial flutter, LBBB—left bundle branch block, PVC—premature ventricular contraction, RBBB—right bundle branch block.

### Table S5: SKAT-O burden test analysis (P/LP variants)

| Phenotype | All genes (n=123) | *ANK2* (n=16) | *KCNE1* (n=9) | *KCNH2* (n=7) | *KCNQ1* (n=30) | *LMNA* (n=7) | *RYR2* (n=31) | *SCN5A* (n=25) |
|---|---|---|---|---|---|---|---|---|
| **Extreme** | | | | | | | | |
| LQTS | **2.2e-10*** | 1 | **5.5e-8*** | **2.2e-10*** | **9.1e-7*** | 1 | 0.9 | **6.5e-5*** |
| SCD | 0.39 | 0.76 | 1 | 1 | 0.84 | 1 | 0.64 | 0.89 |
| VT/VF | **7.7e-5*** | **2.3e-4*** | 0.75 | 1 | 0.45 | **2.2e-10*** | 0.31 | 0.06 |
| **Severe** | | | | | | | | |
| Advanced AV block | **5.4e-7*** | **2.2e-10*** | 0.78 | 1 | 0.64 | **2.9e-7*** | **1.5e-5*** | 0.78 |
| CIEDs | 0.11 | 0.78 | 0.81 | 1 | 0.66 | **2.4e-10*** | 0.06 | 0.81 |
| Sick Sinus | **0.02** | 0.76 | 0.75 | 1 | 0.08 | **2.2e-7*** | 0.09 | 0.74 |
| **Moderate** | | | | | | | | |
| AF/AFL | 0.71 | 0.60 | 0.87 | 0.78 | 0.65 | **7.0e-4*** | 0.21 | 0.68 |
| **Mild** | | | | | | | | |
| LBBB | 0.24 | 0.85 | 0.78 | 1 | 0.58 | **7.4e-7*** | 0.35 | **0.008** |
| PVCs | 0.08 | 0.38 | 0.51 | 0.84 | 0.68 | **0.003** | 0.24 | 0.24 |
| RBBB | 0.80 | 0.01 | 0.72 | 1 | 0.10 | 1 | 0.48 | 0.71 |
| Syncope | 0.10 | 0.19 | 0.41 | 0.66 | 0.20 | **0.03** | 1 | **3.2e-4*** |

SKAT-O analysis (P/LP heterozygotes vs. non-carrier) adjusted for sex, 10 principal components of ancestry, age, and site. Entries indicate p-values. *CACNA1C*, *KCNE2* and *KCNJ2* had 4, 1, and 0 individuals with pathogenic/likely pathogenic variants, respectively, all without any arrhythmia codes, so odds ratios and p-values were not calculated. Bold: Nominal p-value <0.05, unadjusted for multiple testing. * indicates false discovery rate < 0.1 (adjusted for p-values in this table). Abbreviations: LQTS— long QT syndrome, SCD— sudden cardiac death, VT/VF— ventricular tachycardia/ventricular fibrillation, CIED—cardiovascular implantable electronic devices, AF/AFL—atrial fibrillation/atrial flutter, LBBB—left bundle branch block, PVC— premature ventricular contraction, RBBB—right bundle branch block.

**Table S6. SKAT-O burden test analysis (VUS)**

| Phenotype | All genes (n=1838) | *ANK2* (n=493) | *CACNA1C* (n=219) | *KCNE1* (n=23) | *KCNH2* (n=177) | *KCNJ2* (n=27) | *KCNQ1* (n=109) | *LMNA* (n=80) | *RYR2* (n=528) | *SCN5A* (n=284) |
|---|---|---|---|---|---|---|---|---|---|---|
| **Extreme** | | | | | | | | | | |
| LQTS | 0.13 | 0.44 | 0.68 | 0.84 | 0.16 | 0.82 | 0.58 | **3.7e-4*** | 0.41 | 0.26 |
| SCD | 0.62 | 0.79 | 0.89 | 0.90 | 0.48 | 0.88 | **0.02** | 0.73 | 0.64 | 1 |
| VT/VF | 0.44 | 0.23 | 0.72 | 0.76 | 0.06 | 0.72 | 0.68 | **1.4e-9*** | 0.45 | 0.80 |
| **Severe** | | | | | | | | | | |
| Advanced AV block | 0.69 | 0.41 | 0.27 | **0.003** | 1 | **0.02** | 0.78 | **6.1e-9*** | 0.62 | 0.79 |
| CIEDs | 0.79 | **0.49** | 0.53 | 0.84 | 0.55 | **0.02** | 0.28 | **5.7e-8*** | 0.76 | 0.73 |
| Sick Sinus | 0.5 | 0.23 | 0.60 | **0.01** | 1 | 0.06 | 0.03 | 0.85 | 0.82 | 0.48 |
| **Moderate** | | | | | | | | | | |
| AF/AFL | 0.23 | 1 | 0.42 | 0.09 | 0.84 | 0.62 | 0.29 | **0.01** | 0.14 | 0.06 |
| **Mild** | | | | | | | | | | |
| LBBB | 0.69 | 0.72 | 0.87 | 0.81 | 0.62 | 0.70 | 0.56 | **0.005** | 0.15 | 1 |
| PVCs | 0.57 | 1 | 0.35 | 0.06 | 0.45 | 0.38 | 0.48 | 0.08 | 0.51 | 0.66 |
| RBBB | 0.19 | 0.11 | 1 | **0.03** | 0.71 | 0.61 | 0.25 | **3.2e-6*** | 0.44 | 0.73 |
| Syncope | 0.48 | 0.32 | 0.37 | 0.06 | 1 | 0.80 | 0.42 | 0.31 | 0.54 | 0.66 |

SKAT-O analysis (ultra-rare VUS heterozygotes vs. non-carrier) adjusted for sex, 10 principal components of ancestry, age, and site. Values indicate p-values. There were only 3 *KCNE2* VUS heterozygotes so this gene was excluded from the table. Bold: Nominal p-value <0.05, unadjusted for multiple testing. * indicates false discovery rate < 0.1 (adjusted for p-values in this table). Abbreviations: LQTS— long QT syndrome, SCD— sudden cardiac death, VT/VF— ventricular tachycardia/ventricular fibrillation, CIED—cardiovascular implantable electronic devices, AF/AFL—atrial fibrillation/atrial flutter, LBBB—left bundle branch block, PVC— premature ventricular contraction, RBBB—right bundle branch block.

**Table S7: Pre-2017 associations between P/LP heterozygotes and EHR code-derived phenotypes**

| Phenotype | All genes (n=123) | *ANK2* (n=16) | *KCNE1* (n=9) | *KCNH2* (n=7) | *KCNQ1* (n=30) | *LMNA* (n=7) | *RYR2* (n=31) | *SCN5A* (n=25) |
|---|---|---|---|---|---|---|---|---|
| **Extreme** | | | | | | | | |
| LQTS | **8.9 (4.6e-6)*** | 0 (1) | **20.2 (0.006)*** | **17.4 (0.01)*** | **17.2 (7.7e-6)*** | 0 (1) | 0 (1) | 0 (1) |
| SCD | 0 (1) | 0 (1) | 0 (1) | 0 (1) | 0 (1) | 0 (1) | 0 (1) | 0 (1) |
| VT/VF | 2.5 (0.08) | 8.4 (0.06) | 0 (1) | 0 (1) | 0 (1) | **34.4 (4.8e-4)*** | **8.0 (0.01)*** | 0 (1) |
| **Severe** | | | | | | | | |
| Advanced AV block | 2.7 (0.07) | 7.7 (0.10) | 0 (1) | 0 (1) | 0 (1) | **15.2 (0.02)*** | **6.1 (0.02)*** | 3.4 (0.25) |
| CIEDs | 1.8 (0.36) | 0 (1) | 0 (1) | 0 (1) | 0 (1) | **14.6 (0.03)*** | **5.2 (0.05)** | 0 (1) |
| Sick Sinus | **3.6 (0.005)*** | 0 (1) | 0 (1) | 0 (1) | 2.4 (0.40) | **41.3 (7.2e-5)*** | **5.4 (0.05)** | 2.7 (0.36) |
| **Moderate** | | | | | | | | |
| AF/AFL | 1.2 (0.64) | 0 (1) | 0.9 (0.95) | 0 (1) | 0.4 (0.34) | **25.7 (0.002)*** | 2.0 (0.31) | 1.1 (0.95) |
| **Mild** | | | | | | | | |
| LBBB | 2.0 (0.18) | 0 (1) | 0 (1) | 0 (1) | 0 (1) | **36.2 (4.1e-4)*** | 2.5 (0.40) | 2.6 (0.37) |
| PVCs | 1.4 (0.39) | 3.1 (0.32) | 0 (1) | 0 (1) | 0 (1) | 6.4 (0.11) | **4.5 (0.01)*** | 2.2 (0.30) |
| RBBB | 2.0 (0.20) | 7.1 (0.09) | 0 (1) | 0 (1) | 1.7 (0.62) | **13.2 (0.025)*** | 1.9 (0.53) | 2.5 (0.38) |
| Syncope | **1.8 (0.03)*** | 2.0 (0.41) | 2.3 (0.33) | 1.3 (0.83) | 0.3 (0.24) | **13.0 (0.002)*** | 0.8 (0.75) | **6.2 (9.5e-5)*** |

Contingency analysis of Table 2 only using data before 2017 (the year of the beginning of the return of variant results). Similar results were seen to the analysis from the entire dataset (Table 1, S1). Logistic regression adjusted for sex, 10 principal components of ancestry, age, and site. Entries indicate Odds Ratio (p-value). *CACNA1C*, *KCNE2* and *KCNJ2* had 4, 1, and 0 individuals with pathogenic/likely pathogenic variants, respectively, all without any arrhythmia codes, so odds ratios and p-values were not calculated. Bold: Nominal p-value <0.05, unadjusted for multiple testing. * indicates false discovery rate < 0.1 (adjusted for p-values in this table). Abbreviations: LQTS— long QT syndrome, SCD— sudden cardiac death, VT/VF— ventricular tachycardia/ventricular fibrillation, CIED—cardiovascular implantable electronic devices, AF/AFL—atrial fibrillation/atrial flutter, LBBB—left bundle branch block, PVC—premature ventricular contraction, RBBB—right bundle branch block.

**Table S8: Associations between P/LP heterozygotes and EHR code-derived phenotypes in self-reported White participants**

| Phenotype | All genes (n=80) | *ANK2* (n=10) | *KCNE1* (n=8) | *KCNH2* (n=5) | *KCNQ1* (n=20) | *LMNA* (n=4) | *RYR2* (n=14) | *SCN5A* (n=18) |
|---|---|---|---|---|---|---|---|---|
| **Extreme** | | | | | | | | |
| LQTS | **13.8 (2.2e-12)*** | 0 (1) | **28.7 (1.0e-4)*** | **40.7 (4.2e-4)*** | **23.0 (4.4e-8)*** | 0 (1) | 0 (1) | 5.3 (0.12) |
| SCD | 0 (1) | 0 (1) | 0 (1) | 0 (1) | 0 (1) | 0 (1) | 0 (1) | 0 (1) |
| VT/VF | **3.0 (0.02)*** | **12.7 (0.05)** | 0 (1) | 0 (1) | 0 (1) | **79.7 (0.01)*** | **7.1 (0.02)*** | **5.3 (0.04)** |
| **Severe** | | | | | | | | |
| Advanced AV block | 1.7 (0.49) | 0 (1) | 0 (1) | 0 (1) | 0 (1) | 0 (1) | 4.1 (0.21) | 3.8 (0.21) |
| CIEDs | 1.6 (0.52) | 0 (1) | 0 (1) | 0 (1) | 0 (1) | 0 (1) | **8.9 (0.009)*** | 0 (1) |
| Sick Sinus | 2.5 (0.10) | 0 (1) | 0 (1) | 0 (1) | 2.5 (0.39) | 0 (1) | **6.7 (0.03)** | 2.5 (0.41) |
| **Moderate** | | | | | | | | |
| AF/AFL | 1.0 (0.93) | 0 (1) | 0.9 (0.90) | 0 (1) | 0.4 (0.35) | 29.1 (0.08) | 2.3 (0.27) | 1.0 (0.96) |
| **Mild** | | | | | | | | |
| LBBB | 1.2 (0.77) | 0 (1) | 0 (1) | 0 (1) | 0 (1) | 0 (1) | 3.2 (0.29) | 2.7 (0.34) |
| PVCs | 1.4 (0.40) | 3.9 (0.25) | 0 (1) | 0 (1) | 0 (1) | 0 (1) | **5.5 (0.01)*** | 3.4 (0.08) |
| RBBB | 0.4 (0.40) | 12.4 (0.05) | 0 (1) | 0 (1) | 0 (1) | 0 (1) | 1.9 (0.54) | 0 (1) |
| Syncope | **2.1 (0.02)*** | 3.8 (0.14) | 3.0 (0.21) | 2.5 (0.43) | 0.4 (0.43) | 6.1 (0.14) | 1.5 (0.63) | **5.7 (0.001)*** |

Logistic regression adjusted for sex, 10 principal components of genetic ancestry, age, and site. Entries indicate Odds Ratio (p-value). *CACNA1C*, *KCNE2* and *KCNJ2* had 4, 1, and 0 individuals with pathogenic/likely pathogenic variants, respectively, all without any arrhythmia codes, so odds ratios and p-values were not calculated. Bold: Nominal p-value <0.05, unadjusted for multiple testing. * indicates false discovery rate < 0.1 (adjusted for p-values in this table). Abbreviations: LQTS— long QT syndrome, SCD— sudden cardiac death, VT/VF— ventricular tachycardia/ventricular fibrillation, CIED—cardiovascular implantable electronic devices, AF/AFL—atrial fibrillation/atrial flutter, LBBB—left bundle branch block, PVC—premature ventricular contraction, RBBB—right bundle branch block.

**Table S9: Differences in ECG intervals obtained from the EHR as a function of P/LP status**

| Gene | PR (ms) | | | QRS (ms) | | | QTc (ms) | | |
|---|---|---|---|---|---|---|---|---|---|
| | Non-carrier | P/LP carrier | P-value | Non-carrier | P/LP carrier | P-value | Non-carrier | P/LP carrier | P-value |
| No variant (n=7,046) | 160±0.3 | N/A | N/A | 93±0.2 | N/A | N/A | 432±0.3 | N/A | N/A |
| All (n=39) | | **167±5** | **0.04** | | 95±3 | 0.15 | | **451±5** | **7.2e-7*** |
| *KCNE1* (n=5) | | 177±15 | 0.10 | | 87±2 | 0.62 | | 452±12 | 0.10 |
| *KCNH2* (n=5) | | 154±7 | 0.92 | | 79±2 | 0.23 | | **464±17** | **0.003*** |
| *KCNQ1* (n=13) | | 158±6 | 0.62 | | 92±4 | 0.85 | | **455±8** | **0.001*** |
| *LMNA* (n=3) | | 151±15 | 0.44 | | 112±25 | 0.06 | | **459±28** | **0.04** |
| *RYR2* (n=5) | | 158±8 | 0.99 | | 94±5 | 0.90 | | 441±15 | 0.45 |
| *SCN5A* (n=7) | | **190±19** | **4.4e-4*** | | **112±12** | **0.001*** | | 446±15 | 0.12 |

Mean ± standard error of electrocardiographic intervals across P/LP heterozygotes and non-carriers. All intervals (PR, QRS, and QTc) are in milliseconds. P-values are derived from a linear regression adjusted for sex, 10 principal components of ancestry, age, and site. Bold: nominal p-value below 0.05. *False discovery rate <0.1 (adjusted for all p-values in this table). No carriers of P/LP variants in *ANK2*, *KCNE2*, and *KCNJ2*, and only 1 P/LP carrier of *CACNA1C* had ECG data available, so coefficients and p-values were not calculated for these genes. This dataset is plotted in Figure 2A-C.

**Table S10: Differences in ECG intervals obtained from the EHR as a function of VUS status**

| Gene | PR (ms) | | | QRS (ms) | | | QTc (ms) | | |
|---|---|---|---|---|---|---|---|---|---|
| | **Non-carrier** | **VUS carrier** | **P-value** | **Non-carrier** | **VUS carrier** | **P-value** | **Non-carrier** | **VUS carrier** | **P-value** |
| No variant (n=7,046) | 160±0.3 | N/A | N/A | 93±0.3 | N/A | N/A | 432±0.3 | N/A | N/A |
| All (n=585) | | 160±1 | 0.92 | | 93±0.8 | 0.68 | | 432±1.1 | 0.68 |
| *ANK2* (n=170) | | 155±1.8 | **0.006** | | 92±1.2 | 0.21 | | 430±1.8 | 0.39 |
| *CACNA1C* (n=68) | | 157±3.1 | 0.17 | | 91±2.5 | 0.31 | | 432±3 | 0.76 |
| *KCNE1* (n=6) | | 171±8 | 0.53 | | 91±5.2 | 0.39 | | 428±9.3 | 0.45 |
| *KCNH2* (n=48) | | 169±3.8 | **0.04** | | 90±1.6 | 0.19 | | 432±4 | 0.45 |
| *KCNJ2* (n=8) | | 150±6.3 | 0.17 | | 91±2 | 0.66 | | 429±13.4 | 0.68 |
| *KCNQ1* (n=35) | | 161±3.3 | 0.67 | | 94±4.2 | 0.42 | | 441±5.4 | **0.04** |
| *LMNA* (n=32) | | 166±5 | 0.16 | | 100±5.3 | **0.005** | | 442±6.1 | 0.07 |
| *RYR2* (n=155) | | 161±1.9 | 0.88 | | 93±1.4 | 0.95 | | 432±2 | 0.98 |
| *SCN5A* (n=81) | | 166±3.2 | **0.02** | | 94±1.9 | 0.47 | | 431±2.8 | 0.56 |

Mean ± standard error of electrocardiographic intervals across VUS heterozygotes and non-carriers. All intervals (PR, QRS, and QTc) are in milliseconds. P-values are derived from a linear regression adjusted for sex, 10 principal components of ancestry, age, and site. Bold: nominal p-value below 0.05. No association had a false discovery rate <0.1 (adjusted for all p-values in this table). Only 1 P/LP carrier of *KCNE2* had ECG data available, so coefficients and p-values were not calculated for this gene.

**Table S11: Participants with P/LP variants diagnosed with inherited arrhythmia syndromes**

| Age /Sex | Race | Gene | Variant | Class | Diagnosis | Diagnosis Post-ROR? | QTc (ms) | Other documented phenotypes |
|---|---|---|---|---|---|---|---|---|
| 63F | W | *KCNE1* | p.Asp76Asn | P | LQTS | no | 520 | N/A |
| 59F | W | *KCNE1* | p.Arg98Trp | P | LQTS | yes | 515 | Syncope, PVCs |
| 74F | W | *KCNH2* | p.Thr152fs | LP | LQTS | no | 553 | Syncope |
| 59F | W | *KCNH2* | p.Gly921fs | LP | LQTS | yes | 510 | N/A |
| 48F | W | *KCNH2* | p.Ile82Thr | LP | LQTS | yes | 468 | N/A |
| 66F | N/A | *KCNH2* | c.308-2A>G | LP | LQTS | yes | 506 | N/A |
| 15F | B | *KCNQ1* | p.Leu142Argfs | LP | LQTS | no | 441 | N/A |
| 57F | W | *KCNQ1* | p.Phe340del | P | LQTS | no | 534 | N/A |
| 65F | W | *KCNQ1* | p.Tyr315Cys | LP | LQTS | no | 490 | N/A |
| 10F | B | *KCNQ1* | p.Gly186Asp | LP | LQTS | yes | 471 | N/A |
| 67M | W | *KCNQ1* | p.Arg632Glnfs | P | LQTS | yes | 436 | N/A |
| 67F | W | *KCNQ1* | p.Arg518Ter | P | LQTS | yes | 517 | N/A |
| 74F | W | *KCNQ1* | Exon 4-7 del. | P | LQTS | yes | 488 | N/A |
| 76M | W | *KCNQ1* | p.Arg192Cysfs | P | LQTS | yes | 466 | LAFB |
| 62M | W | *LMNA* | p.Arg377Cys | LP | DCM | no | 594 | AF, QRS 166ms, VT |
| 74F | W | *SCN5A* | p.Arg535Ter | P | BrS | no | 428(QT) | LBBB, Syncope, PVCs |
| 18M | W | *SCN5A* | c.4245+1G>C | LP | BrS | yes | 376 | Type 2 Brugada Pattern |
| 69F | W | *SCN5A* | p.Gly1319Val | P | LQTS | yes | 439 | Syncope |

All variants were heterozygous. Abbreviations: LQTS—Long QT syndrome, DCM—Dilated cardiomyopathy, BrS—Brugada syndrome, PVC—premature ventricular contraction, AF—atrial fibrillation, VT—ventricular tachycardia, LBBB—left bundle branch block, W—White, B—Black, P—pathogenic, LP—likely pathogenic. The maximum available QTc interval (QT interval corrected for heart rate using Bazett's formula) is displayed in the QTc column. For the individual with the *SCN5A* p.Arg535Ter variant, only QT, but not QTc, was available.

**Table S12: Changes in clinical management in participants with returned P/LP variants**

| Action | All (n=51) | LQTS (n=15) | *LMNA*-DCM (n=1) | BrS (n=2) | No Dx (n=33) |
|---|---|---|---|---|---|
| Seen by specialist | 30 | 12 | 1 | 2 | 15 |
| Electrocardiogram | 38 | 15 | 1 | 2 | 20 |
| Echocardiogram | 23 | 10 | 1 | 1 | 11 |
| New diagnosis made | 11 | 10 | 0 | 1 | 0 |
| Drug change | 6 | 5 | 0 | 0 | 1 |
| Medication counseling | 3 | 3 | 0 | 0 | 0 |
| ICD implanted | 1 | 0 | 0 | 1 | 0 |

DCM—Dilated Cardiomyopathy, LQTS—Long QT syndrome, BrS—Brugada syndrome, Dx—Diagnosis, ICD—Implantable Cardioverter-Defibrillator. Electrocardiograms and echocardiograms were from either before or after return of results; all other changes were restricted to after RoR actions.

### Table S13: Additional properties of variants studied *in vitro*

| Variant | gnomAD AF (x10^-5) | REVEL | Hom. # cells | Hom Peak Mean (SE) | Het. # cells | Het. Peak Mean (SE) | ACMG Criteria | ACMG Class. |
|---|---|---|---|---|---|---|---|---|
| KCNH2 p.Ala244Val | 0 | 0.257 | 32 | 98.6 (5.1) | — | — | BS3 BP4 PM2 PP2 | VUS→LB |
| KCNH2 p.Gly924Trp | 1.3 | 0.586 | 38 | 78.8 (5.4) | — | — | BS3 PM1m PP2 | VUS |
| KCNH2 p.Pro1132Ala | 1.8 | 0.54 | 35 | 83.0 (4.6) | — | — | BS3 PM1m PP2 | VUS |
| KCNH2 p.Pro1075Leu | 2 | 0.788 | 21 | 86.6 (6.5) | — | — | BS3 PM1m PP2 | VUS |
| KCNH2 p.Glu682Asp | 0 | 0.813 | 37 | 81.5 (6.0) | — | — | BS3 PM1m PM2 PP2 | VUS |
| KCNH2 p.Cys1117Ser | 0 | 0.569 | 21 | 82.7 (8.4) | — | — | BS3 PM1m PM2 PP2 | VUS |
| KCNQ1 p.Gly626Ser | 1.1 | 0.566 | 63 | 88.6 (8.8) | 37 | 115 (13.9) | BS3 BP4 PM1p PP2 | VUS→LB |
| KCNQ1 p.Arg181Leu | 0 | 0.895 | 44 | 99.5 (9.4) | 31 | 99.2 (14.7) | BS3 PM1m PM2 PP2 PP3 | VUS→LP |
| KCNQ1 p.Cys381Trp | 0 | 0.798 | 54 | 73.5 (7) | 91 | 103.7 (8.1) | PM1m PM2 PP2 PP3 | VUS→LP |
| KCNQ1 p.Arg533Trp | 0 | 0.77 | 31 | 7.4 (1.2) | 44 | 43.6 (5.4) | PS3m PM1m PM2 PP2 PP3 | VUS→LP |
| KCNQ1 p.Val280Glu | 0 | 0.944 | 29 | 3.5 (0.9) | 30 | 26.3 (4.8) | PS3 PM1m PM2 PP2 PP3 | VUS→P |
| KCNQ1 p.Gly186Asp | 0 | 0.951 | 36 | 6.5 (1.2) | 31 | 31.6 (6.1) | PS3 PM1m PM2 PP2 PP3 PP5 | LP→P |
| KCNQ1 p.Ala31Val | 0 | 0.32 | 31 | 119 (14.9) | — | — | BS3 PM2 PP2 | VUS |
| KCNQ1 p.Gly57Ser | 0 | 0.349 | 43 | 94.8 (8.8) | — | — | BS3 PM2 PP2 | VUS |
| KCNQ1 p.Arg181His | 0 | 0.841 | 45 | 111.3 (8.9) | 44 | 103.7 (11.6) | BS3 PM1m PM2 PP2 | VUS |
| KCNQ1 p.Ser475Asn | 0 | 0.525 | 39 | 81.8 (11.3) | — | — | BS3 PM1p PM2 PP2 | VUS |
| KCNQ1 p.Leu491Val | 0 | 0.68 | 46 | 76 (6.2) | — | — | BS3 PM1p PM2 PP2 | VUS |
| KCNQ1 p.Thr495Pro | 0 | 0.644 | 48 | 77 (6.7) | 41 | 81.6 (9.7) | BS3 PM1p PM2 PP2 | VUS |
| KCNQ1 p.Glu449Lys | 2.5 | 0.624 | 43 | 107.1 (10.5) | — | — | BS3 PM1p PP2 | VUS |
| KCNQ1 p.Gly57Arg | 0 | 0.389 | 38 | 113 (10.1) | — | — | BS3 PM2 PP2 | VUS |
| KCNQ1 p.Asp388Asn | 0.8 | 0.568 | 51 | 103.1 (9.4) | — | — | BS3 PM1m PM2 PP2 | VUS |
| KCNQ1 p.Glu290Lys | 1.8 | 0.683 | 44 | 57.5 (6.9) | 73 | 101.9 (10.1) | PM1m PP2 | VUS |
| KCNQ1 p.Pro448Leu | 2.5 | 0.592 | 37 | 49.4 (6.7) | 38 | 119.1 (10.5) | PM1p PP2 | VUS |
| KCNQ1 p.Ser66Phe | 0 | 0.649 | 49 | 70.5 (8.9) | 37 | 87.5 (12.2) | PM2 PP2 | VUS |
| KCNQ1 p.Ala287Val | 0 | 0.741 | 41 | 102.5 (11.3) | 56 | 252.6 (24) | PM1m PM2 PP2 | VUS |
| KCNQ1 p.Pro441Ser | 2.5 | 0.643 | 39 | 38.7 (6) | 92 | 84.9 (5.6) | PM1p PP2 PP3 | VUS |
| KCNQ1 p.Arg452Leu | 0.8 | 0.6 | 47 | 65.1 (7.1) | 51 | 63.7 (7) | PM1p PM2 PP2 | VUS |

| | | | | | | | |
|---|---|---|---|---|---|---|---|
| KCNQ1 p.Gln531Glu | 0 | 0.661 | 38 | 49.1 (7.5) | 43 | 82.5 (9) | PM1m PM2 PP2 | VUS |
| KCNQ1 p.Lys398Arg | 0 | 0.653 | 54 | 60.9 (7.3) | 51 | 99 (11.7) | PM1p PM2 PP2 | VUS |
| KCNQ1 p.Asp488Glu | 0.8 | 0.653 | 47 | 69.6 (7) | 51 | 80.6 (10.2) | PM1p PM2 PP2 | VUS |
| KCNQ1 p.Gly635Arg | 1.8 | 0.518 | 44 | 70.5 (7.6) | 67 | 75.1 (7.8) | PM1p PP2 | VUS |
| KCNQ1 p.Gly57Val | 0 | 0.373 | 45 | 137.2 (10.7) | — | — | PM2 PP2 | VUS |
| KCNQ1 p.Pro67Arg | 0 | 0.388 | 36 | 97.2 (12.3) | 36 | 150.1 (22.6) | PM2 PP2 | VUS |
| KCNQ1 p.Thr444Met | 1.4 | 0.684 | 42 | 70.3 (7.4) | 92 | 88 (7.7) | PM1p PP2 PP3 | VUS |
| KCNQ1 p.Ile574Val | 0.4 | 0.596 | 56 | 93.6 (8.3) | 26 | 133 (20.1) | BP4 PM1m PM2 PP2 | VUS |
| KCNQ1 p.Arg519His | 2.5 | 0.876 | 33 | 16.4 (4.1) | 39 | 50.5 (5.6) | PS3m PM1m PP2 | VUS |
| KCNQ1 p.Asp649Asn | 2.3 | 0.404 | 46 | 45.8 (5.2) | 69 | 120.4 (12.5) | BP4 BP6 PM1p PP2 | LB |
| SCN5A p.Pro1123Leu | 0 | 0.751 | 55 | 106.0 (8.1) | — | — | BS3 BP4 PM2 PP2 | VUS→LB |
| SCN5A p.Phe1794Ile | 0 | 0.972 | 56 | 16.0 (2.7) | — | — | PS3 PM1p PM2 PP2 PP3 | VUS→LP |
| SCN5A p.Gly1605Asp | 0.4 | 0.952 | 39 | 145.1 (15.2) | — | — | PS3 PM1m PM2 PP2 | VUS→LP |
| SCN5A p.Asp1741Asn | 1.2 | 0.33 | 23 | 157.8 (24.3) | — | — | BP4 PS3 PM1m PP2 | VUS→LP |
| SCN5A p.Ile848Phe | 0.4 | 0.961 | 70 | 70.9 (5.1) | — | — | PM1m PM2 PP2 PP3 | VUS→LP |
| SCN5A p.Ala662Ser | 2.1 | 0.834 | 67 | 87.4 (6.9) | — | — | BS3 PP2 | VUS |
| SCN5A p.Gly552Trp | 1.4 | 0.635 | 45 | 107.2 (8.4) | — | — | BS3 PP2 PP3 | VUS |
| SCN5A p.Lys1500Asn | 0 | 0.722 | 39 | 78.2 (6.3) | — | — | BS3 PM2 PP2 PP3 | VUS |
| SCN5A p.Ser835Ala | 1.1 | 0.906 | 59 | 102.6 (6.2) | — | — | BS3 PM1m PP2 PP3 | VUS |
| SCN5A p.Tyr1228His | 0.4 | 0.854 | 68 | 114.8 (7.8) | — | — | BS3 PM1m PM2 PP2 | VUS |
| SCN5A p.Ser172Thr | 0 | 0.863 | 74 | 109.3 (8.6) | — | — | BS3 PM1m PM2 PP2 | VUS |
| SCN5A p.Gly1084Ser | 0.8 | 0.557 | 36 | 101.7 (9.5) | — | — | BS3 PM2 PP2 | VUS |
| SCN5A p.Asn291His | 1.1 | 0.586 | 55 | 82.6 (10.0) | — | — | BS3 PM1m PP2 | VUS |
| SCN5A p.Ile94Val | 1.2 | 0.313 | 23 | 120.4 (14.9) | — | — | BS3 PM1m PP2 | VUS |
| SCN5A p.Gln245Lys | 0.8 | 0.906 | 34 | 116.4 (9.5) | — | — | BS3 PM1p PM2 PP2 | VUS |

The PS3 criterion was applied at the strong (PS3) or moderate (PS3m) level. The PM1 criterion was applied at the moderate (PM1) or supporting (PM1p) level.

### Table S14: Mutagenesis primers

| Gene | Variant | Direction | Primer |
|---|---|---|---|
| *SCN5A* | p.Ala662Ser | F | CACGGCAGCGGTCCCTCAGCGCA |
| *SCN5A* | p.Gly552Trp | F | AAAACAGCACAGCGTGGGAGAGCGAGAGC |
| *SCN5A* | p.Pro1123Leu | F | GGAAAGCGGAACTCCAGGCCCCAGG |
| *SCN5A* | p.Ser835Ala | F | AGATCATCGGGAACGCAGTGGGGGCACTG |
| *SCN5A* | p.Phe1794Ile | F | AGGACGACTTCGATATGATCTATGAGATCTGGGAG |
| *SCN5A* | p.Gly1605Asp | F | CCTCTCCATCGTGGACACTGTGCTCTCGG |
| *SCN5A* | p.Asp1741Asn | F | GCTCTCGGGGGAACTGCGGGAGC |
| *SCN5A* | p.Tyr1228His | F | CCTTCGAGGACATCCACCTAGAGGAGCGG |
| *SCN5A* | p.Ser172Thr | F | ATTTACACCTTTGAGACTCTGGTCAAGATTC |
| *SCN5A* | p.Gly1084Ser | F | CAGCCTGTGTCCAGTGGCCCAGAGG |
| *SCN5A* | p.Lys1500Asn | F | AGTACTACAATGCCATGAAGAACCTGGGCTCCAAG |
| *SCN5A* | p.Asn291His | F | CTCAACGGCACCCACGGCTCCGTGG |
| *SCN5A* | p.Ile848Phe | F | TGACACTGGTGCTTGCCTTCATCGTGTTCATCTTT |
| *SCN5A* | p.Ile94Val | F | ATAGCACCCAAAAGACTTTCGTCGTACTGAATAAAGGCAAG |
| *SCN5A* | p.Gln245Lys | F | GCAGCGCGCTGATCAAGTCTGTGAAGAAGC |
| *KCNH2* | p.Gly924Trp | F | GGCCGGCCGTGGGGGCCGTG |
| *KCNH2* | p.Ala224Val | F | CCCCGCAGCGTGCCCGGCCAG |
| *KCNH2* | p.Pro1132Ala | F | CCCCAAGAAGGCGCCACACGACGCC |
| *KCNH2* | p.Pro1075Leu | F | TGACGCTGGTCCTGCCCGCCTACAG |
| *KCNH2* | p.Glu682Asp | F | GCGGGTGCGGGACTTCATCCGCTTC |
| *KCNH2* | p.Cys1117Ser | F | CCAGTTCATGGCGAGTGAGGAGCTGCC |
| *KCNQ1* | p.Ala31Val | F | CCTGGTCAAGAAGTGCCCCTTCTCGCTGG |
| *KCNQ1* | p.Gly57Ser | F | TCGCGCCCAGCGCCCCAGGTCCCGCGC |
| *KCNQ1* | p.Gly57Arg | F | TCGCGCCCCGCGCCCCAGGTCCCGCGC |
| *KCNQ1* | p.Gly57Val | F | CGCGCCCGTCGCCCCAGGTCCCGCGCC |
| *KCNQ1* | p.Ser66Phe | F | GCGCCCCCTGCGTTCCCGGCC |
| *KCNQ1* | p.Pro67Arg | F | CCTGCGTCCCGGGCCGCGCCC |
| *KCNQ1* | p.Arg181His | F | GCTGCCACAGCAAGTACGTGGGCCTCTGG |
| *KCNQ1* | p.Arg181Leu | F | GCTGCCTCAGCAAGTACGTGGGCCTCTGG |
| *KCNQ1* | p.Gly186Asp | F | AGTACGTGGACCTCTGGGGGCGGCTGCG |
| *KCNQ1* | p.Val280Glu | F | TCGTACTTTGAGTACCTGGCTGAGAAGGACGCG |
| *KCNQ1* | p.Ala287Val | F | GGCTGAGAAGGACGTGGTGAACGAGTCAG |
| *KCNQ1* | p.Glu290Lys | F | CGGTGAACAAGTCAGGCCGCGTGGAGTTCG |
| *KCNQ1* | p.Cys381Trp | F | ATGGAGGTGGTATGCTGCCGAGAACCCCGACTC |
| *KCNQ1* | p.Asp388Asn | F | ACCCCAACTCCTCCACCTGGAAGATCTACATCC |
| *KCNQ1* | p.Lys398Arg | F | TCTACATCCGGAGGGCCCCCCGGAGCCACAC |
| *KCNQ1* | p.Pro441Ser | F | GCTCACAGTCTCCCATATCACGTGCGACCCCC |
| *KCNQ1* | p.Thr444Met | F | CCCCCATATCATGTGCGACCCCCCAGAAGAGCG |
| *KCNQ1* | p.Pro448Leu | F | CGTGCGACCCCCTAGAAGAGCGGCG |
| *KCNQ1* | p.Glu449Lys | F | ACCCCCCAAAAGAGCGGCGGCTGGACCACT |
| *KCNQ1* | p.Arg452Leu | F | CCAGAAGAGCGGCTGCTGGACCACTTC |
| *KCNQ1* | p.Ser475Asn | F | GAAGTGAACATGCCCCATTTCATGAGAACCAACAGC |
| *KCNQ1* | p.Asp488Glu | F | CGCCGAGGAACTGGACCTGGAAGGGGAGACTC |
| *KCNQ1* | p.Leu491Val | F | CTGGACGTGGAAGGGGAGACTCTGCTGACAC |
| *KCNQ1* | p.Thr495Pro | F | AGGGGAGCCTCTGCTGACACCCATCACCCAC |
| *KCNQ1* | p.Arg519His | F | TCGACACATGCAGTACTTTGTGGCCAAGAAGAAATTC |
| *KCNQ1* | p.Gln531Glu | F | AGAAATTCCAGGAAGCGCGGAAGCCTTACGATGTGC |
| *KCNQ1* | p.Arg533Trp | F | CCAGCAAGCGTGGAAGCCTTACGATGTGCGGGAC |
| *KCNQ1* | p.Ile574Val | F | CTGTTCGTCTCCGTCTCAGAAAAGAGCAAGGATCG |
| *KCNQ1* | p.Gly626Ser | F | GCACCCCCAGCAGCGGCGGCCCCCCCA |
| *KCNQ1* | p.Gly635Arg | F | CCCAGAGAGGGCAGGGCCCACATCA |
| *KCNQ1* | p.Asp649Asn | F | TCCGTCAACCCTGAGCTCTTCCTGCCCAG |
| *KCNQ1* | p.Ala31Val | R | GGCACTTCTTGACCAGGCCCGCGCTGCCCC |
| *KCNQ1* | p.Gly57Ser | R | TGGGGCGCTGGGCGCGATGGGCGCGTAG |
| *KCNQ1* | p.Gly57Arg | R | TGGGGCGCGGGGCGCGATGGGCGCGTAG |

| | | | |
|---|---|---|---|
| *KCNQ1* | p.Gly57Val | R | CTGGGGCGACGGGCGCGATGGGCGCGT |
| *KCNQ1* | p.Ser66Phe | R | GGCCGGGAACGCAGGGGGCGC |
| *KCNQ1* | p.Pro67Arg | R | GGGCGCGGCCCGGGACGCAGG |
| *KCNQ1* | p.Arg181His | R | GTACTTGCTGTGGCAGCCGGCGGACCAGAG |
| *KCNQ1* | p.Arg181Leu | R | GTACTTGCTGAGGCAGCCGGCGGACCAGAG |
| *KCNQ1* | p.Gly186Asp | R | CCAGAGGTCCACGTACTTGCTGCGGCAGC |
| *KCNQ1* | p.Val280Glu | R | AGGTACTCAAAGTACGAGGAGAAGATGAGGCCC |
| *KCNQ1* | p.Ala287Val | R | CTGACTCGTTCACCACGTCCTTCTCAGCC |
| *KCNQ1* | p.Glu290Lys | R | GCCTGACTTGTTCACCGCGTCCTTCTCAGCC |
| *KCNQ1* | p.Cys381Tyr | R | GCATAGTACCTCCATGCGGTCTGAATGAGTGAG |
| *KCNQ1* | p.Cys381Trp | R | CAGCATACCACCTCCATGCGGTCTGAATGAGTG |
| *KCNQ1* | p.Asp388Asn | R | GGTGGAGGAGTTGGGGTTCTCGGCAGCATAGCAC |
| *KCNQ1* | p.Lys398Arg | R | GGCCCTCCGGATGTAGATCTTCCAGGTGGAG |
| *KCNQ1* | p.Val416Met | R | TTTCTTTACCACCATAGACTTCTTGGGTTTGGGGCTGGG |
| *KCNQ1* | p.Pro441Ser | R | TATGGGAGACTGTGAGCATCTTCTCTCCAGGAG |
| *KCNQ1* | p.Thr444Met | R | CGCACATGATATGGGGGACTGTGAGCATCTTCTC |
| *KCNQ1* | p.Pro448Leu | R | CGCCGCTCTTCTAGGGGGTCGCACG |
| *KCNQ1* | p.Glu449Lys | R | CCGCTCTTTTGGGGGGTCGCACGTGATATGG |
| *KCNQ1* | p.Glu450Lys | R | CGCCGCTTTTCTGGGGGGTCGCACGTGATATG |
| *KCNQ1* | p.Arg452Leu | R | GAAGTGGTCCAGCAGCCGCTCTTCTGG |
| *KCNQ1* | p.Ser475Asn | R | TGGGGCATGTTCACTTCCAGCAGTGTTGGGCTCTTC |
| *KCNQ1* | p.Asp488Glu | R | GGTCCAGTTCCTCGGCGAAGCTGTTGGTTCTC |
| *KCNQ1* | p.Leu491Val | R | TCCCCTTCCACGTCCAGGTCCTCGGCGAAGCT |
| *KCNQ1* | p.Thr495Pro | R | TCAGCAGAGGCTCCCCTTCCAGGTCCAGGTC |
| *KCNQ1* | p.Arg519His | R | AGTACTGCATGTGTCGAATGACCTTAATGGTGGCCCG |
| *KCNQ1* | p.Gln531Glu | R | CGCTTCCTGGAATTTCTTCTTGGCCACAAAGTACTG |
| *KCNQ1* | p.Arg533Trp | R | GCTTCCACGCTTGCTGGAATTTCTTCTTGGCCAC |
| *KCNQ1* | p.Ile574Val | R | GAGACGGAGACGAACAGTGAGGGCTTCCCAATGGAC |
| *KCNQ1* | p.Gly626Ser | R | GCCGCTGCTGGGGGTGCTGCCACCGTGC |
| *KCNQ1* | p.Gly635Arg | R | TGATGTGGGCCCTGCCCTCTCTGGG |
| *KCNQ1* | p.Asp649Asn | R | AGCTCAGGGTTGACGGAGCCGCCACTGCCG |

*Single primer mutagenesis reactions were used for *SCN5A* and *KCNH2*. F=forward, R=reverse.

**Figure S1: Penetrance of variant carriers by individual variant (*ANK2*, *CACNA1C*, *KCNE2*, *KCNJ2*, and *RYR2*)**

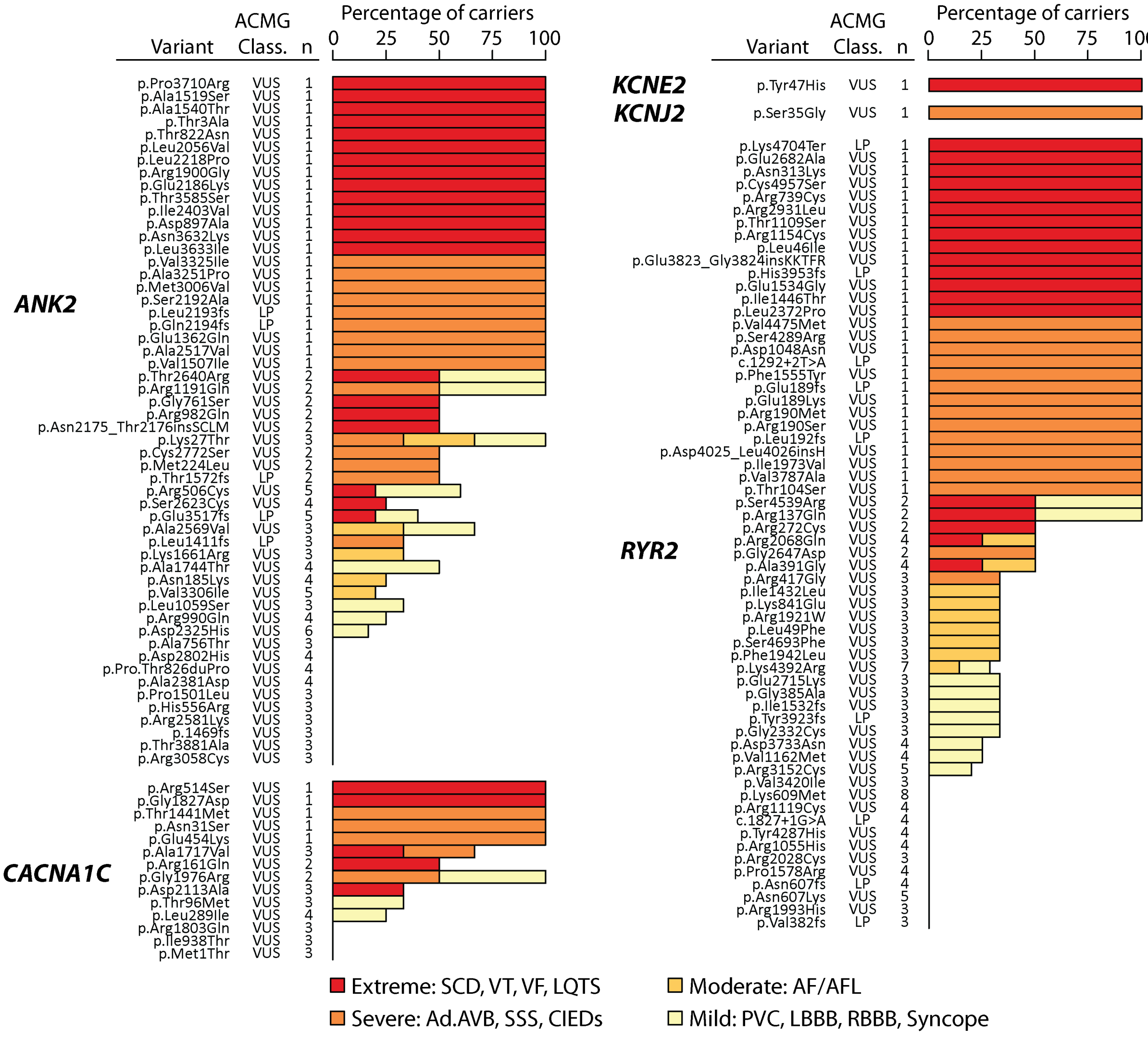


Text columns indicate gene, variant name, number of carriers in study, and variant ACMG classification. Selected variants in *ANK2*, *CACNA1C*, *KCNJ2*, *KCNE2*, and *RYR2* are shown. A similar plot for the remaining genes in this study is presented in Figure 4. Shown variants have a gnomAD minor allele frequency below 2.5e-5 and are either 1) present in at least 3 eMERGE-III heterozygous participants or 2) present in at least 1 heterozygous participant with an extreme or severe phenotype. Abbreviations: SCD: sudden cardiac death, LQTS—long QT syndrome, VT—ventricular tachycardia, VF—ventricular fibrillation, AF/AFL—atrial fibrillation/atrial flutter, PVC—premature ventricular contraction, Ad.AVB—advanced atrioventricular block, SSS—sick sinus syndrome, CIEDs—cardiovascular implantable electronic devices, LBBB—left bundle branch block, RBBB—right bundle branch block, P/LP—pathogenic/likely pathogenic, VUS—variant of uncertain significance.

**Figure S2: *In vitro* functional characterization of *KCNH2* variants**

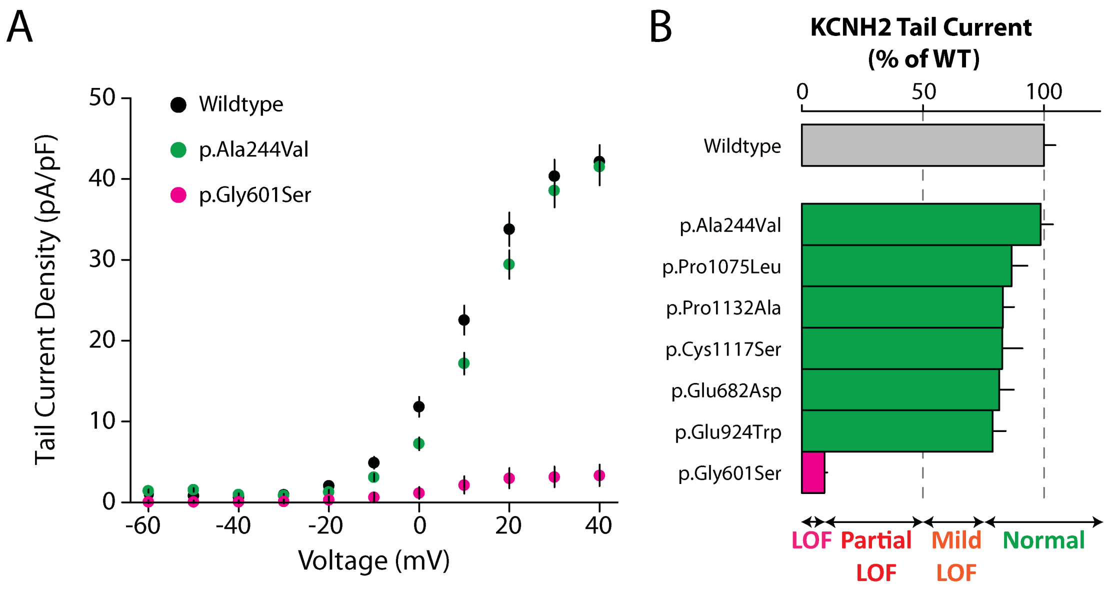


A. Representative tail current-voltage plots for *KCNH2* for selected variants. p.Gly601Ser is a control loss-of-function variant not present in eMERGE-III participants. B. Tail current density was measured as pA/pF and normalized to wildtype. Experiments were performed in Human Embryonic Kidney 293 cells and data was generated using the SyncroPatch instrument. The full patch clamp dataset, including numbers of replicate cells for each variant are listed in File S2.

**File S1: Diagnostic and procedure codes used to generate EHR phenotypes (.docx)**

**File S2: All measured patch clamp parameters for each variant. (.xlsx)**
