## Supplementary material for "Arrhythmia variant associations and reclassifications in the eMERGE-III sequencing study": File S1

**Supplemental File 1. Diagnostic and procedure codes used to generate EHR phenotypes**

**Table 1. Diagnostic and Procedure codes used to identify subjects with atrial fibrillation or atrial flutter from the Electronic Health Records**

| **ICD-9 code** | **Description** |
| --- | --- |
| 427.31 | Atrial fibrillation |
| 427.32 | Atrial flutter |
| **ICD-10 code** | **Description** |
| I48.0 | Paroxysmal atrial fibrillation |
| I48.1 | Persistent atrial fibrillation |
| I48.2 | Chronic atrial fibrillation |
| I48.3 | Typical atrial flutter |
| I48.4 | Atypical atrial flutter |
| I48.9 | Unspecified atrial fibrillation and atrial flutter |
| I48.91 | Unspecified atrial fibrillation |
| I48.92 | Unspecified atrial flutter |
| **CPT code** | **Description** |
| 33254 | Operative tissue ablation and reconstruction of atria, limited (eg, modified maze procedure) |
| 33255 | Operative tissue ablation and reconstruction of atria, extensive (eg, maze procedure); with cardiopulmonary bypass |
| 33256 | Operative tissue ablation and reconstruction of atria, extensive (eg, maze procedure); without cardiopulmonary bypass |
| 33257 | [Operative tissue ablation and reconstruction of atria, performed at the time of other cardiac procedure(s), limited (eg, modified maze procedure) (List separately in addition to code for primary procedure)](https://correctcodechek.decisionhealth.com/Cpt/Search.aspx?st=0&ss=Tabular&sk=33257&vd=07/01/2019) |
| 33258 | [Operative tissue ablation and reconstruction of atria, performed at the time of other cardiac procedure(s), extensive (eg, maze procedure), without cardiopulmonary bypass (List separately in addition to code for primary procedure)](https://correctcodechek.decisionhealth.com/Cpt/Search.aspx?st=0&ss=Tabular&sk=33258&vd=07/01/2019) |
| 33259 | [Operative tissue ablation and reconstruction of atria, performed at the time of other cardiac procedure(s), extensive (eg, maze procedure), with cardiopulmonary bypass (List separately in addition to code for primary procedure)](https://correctcodechek.decisionhealth.com/Cpt/Search.aspx?st=0&ss=Tabular&sk=33259&vd=07/01/2019) |
| 33265 | Endoscopy, surgical; operative tissue ablation and reconstruction of atria, limited (eg, modified maze procedure), without cardiopulmonary bypass |
| 33266 | Endoscopy, surgical; operative tissue ablation and reconstruction of atria, extensive (eg, maze procedure), without cardiopulmonary bypass |
| 93656 | Comprehensive electrophysiologic evaluation including transseptal catheterizations, insertion and repositioning of multiple electrode catheters with induction or attempted induction of an arrhythmia including left or right atrial pacing/recording when necessary, right ventricular pacing/recording when necessary, and His bundle recording when necessary with intracardiac catheter ablation of atrial fibrillation by pulmonary vein isolation |
| 93657 | Additional linear or focal intracardiac catheter ablation of the left or right atrium for treatment of atrial fibrillation remaining after completion of pulmonary vein isolation (List separately in addition to code for primary procedure) |
| 92960 | Cardioversion, elective, electrical conversion of arrhythmia; external |
| 92961 | Cardioversion, elective, electrical conversion of arrhythmia; internal (separate procedure) |

**Table 2. Diagnostic codes used to identify subjects with long QT syndrome from the Electronic Health Records**

| **ICD-9 code** | **Description** |
| --- | --- |
| 426.82 | Long QT syndrome |
| **ICD-10 code** | **Description** |
| I45.81 | Long QT syndrome |

**Table 3. Diagnostic codes used to identify subjects with ventricular tachycardia or ventricular fibrillation (VT/VF) from the Electronic Health Records**

| **ICD-9 code** | **Description** |
| --- | --- |
| 427.42 | Ventricular flutter |
| 427.10 | Paroxysmal ventricular tachycardia |
| 427.41 | Ventricular Fibrillation |
| **ICD-10 code** | **Description** |
| I47.2 | Ventricular tachycardia |
| I49.0 | Ventricular fibrillation and flutter |
| I49.01 | Ventricular fibrillation |
| I49.02 | Ventricular flutter |

**Table 4. Diagnostic codes used to identify subjects with advanced atrioventricular block from the Electronic Health Records**

| **ICD-9 code** | **Description** |
| --- | --- |
| 426.0 | Atrioventricular block, complete |
| 426.0 | Atrioventricular block, complete |
| 426.12 | Mobitz (type) II atrioventricular block |
| 426.13 | Other second degree atrioventricular block |
| 426.51 | Right bundle branch block and left posterior fascicular block |
| 426.52 | Right bundle branch block and left anterior fascicular block |
| 426.54 | Trifascicular block |
| **ICD-10 code** | **Description** |
| I44.1 | Atrioventricular block, second degree |
| I44.2 | Atrioventricular block, complete |
| I45.2 | Bifascicular block |
| I45.3 | Trifascicular block |

**Table 5. Diagnostic codes used to identify subjects with left bundle branch block from the Electronic Health Records**

| **ICD-9 code** | **Description** |
| --- | --- |
| 426.2 | Left bundle branch hemiblock |
| 426.3 | Other left bundle branch block |
| **ICD-10 code** | **Description** |
| I44.7 | Left bundle-branch block, unspecified |

**Table 6. Diagnostic codes used to identify subjects with right bundle branch block from the Electronic Health Records**

| **ICD-9 code** | **Description** |
| --- | --- |
| 426.4 | Right bundle branch block |
| **ICD-10 code** | **Description** |
| I45.10 | Unspecified right bundle-branch block |
| I45.19 | Other right bundle-branch block |

**Table 7. Diagnostic codes used to identify subjects with sick sinus syndrome from the Electronic Health Records**

| **ICD-9 code** | **Description** |
| --- | --- |
| 427.81 | Sinoatrial node dysfunction |
| **ICD-10 code** | **Description** |
| I49.5 | Sick sinus syndrome |

**Table 8. Diagnostic codes used to identify subjects with premature ventricular contractions (PVCs) from the Electronic Health Records**

| **ICD-9 code** | **Description** |
| --- | --- |
| 427.69 | Other premature contraction |
| **ICD-10 code** | **Description** |
| I49.3 | Ventricular premature depolarization |

**Table 9. Diagnostic codes used to identify subjects with syncope from the Electronic Health Records**

| **ICD-9 code** | **Description** |
| --- | --- |
| 780.2 | Syncope and collapse |
| **ICD-10 code** | **Description** |
| R55.9 | Syncope and collapse |

**Table 10. Diagnostic and procedure codes used to identify subjects with sudden arrest, sudden death, and sudden infant death syndrome from the Electronic Health Records**

| **ICD-9 code** | **Description** |
| --- | --- |
| 427.5 | Cardiac arrest |
| 798 | Sudden infant death syndrome |
| 798.1 | Instantaneous death |
| 798.2 | Death occurring in less than 24 hours from onset of symptoms, not otherwise explained |
| 798.9 | Unattended death |
| V12.53 | Personal history of sudden cardiac arrest |
| **ICD-10 code** | **Description** |
| I46.9 | Cardiac arrest |
| P29.81 | Cardiac arrest (newborn) |
| R99 | Death cause unknown |
| Z84.82 | Sudden infant death syndrome |
| Z86.74 | Cardiac arrest (death) successfully resuscitated, personal history |
| **CPT code** | **Description** |
| 92950 | Cardiopulmonary resuscitation |

**Table 11. Procedure codes used to identify subjects with presence of cardiac implantable electronic devices (CIEDs) from the Electronic Health Records**

| **CPT code** | **Description** |
| --- | --- |
| 33200 | Insertion of heart pacemaker |
| [33202](applewebdata://73F254D4-0729-44C2-A445-B1B00C8F2A86/../../../Downloads/%252525250a%2525252509%2525252509%2525252509%2525252509%2525252509%2525252509%2525252509%2525252509%2525252509Detail.aspx%2525253FCode=33202) | Insertion of epicardial electrode(s); open incision (eg, thoracotomy, median sternotomy, subxiphoid approach) |
| [33203](applewebdata://73F254D4-0729-44C2-A445-B1B00C8F2A86/../../../Downloads/%252525250a%2525252509%2525252509%2525252509%2525252509%2525252509%2525252509%2525252509%2525252509%2525252509Detail.aspx%2525253FCode=33203) | Insertion of epicardial electrode(s); endoscopic approach (eg, thoracoscopy, pericardioscopy) |
| [33206](applewebdata://73F254D4-0729-44C2-A445-B1B00C8F2A86/../../../Downloads/%252525250a%2525252509%2525252509%2525252509%2525252509%2525252509%2525252509%2525252509%2525252509%2525252509Detail.aspx%2525253FCode=33206) | Insertion of new or replacement of permanent pacemaker with transvenous electrode(s); atrial |
| [33207](applewebdata://73F254D4-0729-44C2-A445-B1B00C8F2A86/../../../Downloads/%252525250a%2525252509%2525252509%2525252509%2525252509%2525252509%2525252509%2525252509%2525252509%2525252509Detail.aspx%2525253FCode=33207) | Insertion of new or replacement of permanent pacemaker with transvenous electrode(s); ventricular |
| [33208](applewebdata://73F254D4-0729-44C2-A445-B1B00C8F2A86/../../../Downloads/%252525250a%2525252509%2525252509%2525252509%2525252509%2525252509%2525252509%2525252509%2525252509%2525252509Detail.aspx%2525253FCode=33208) | Insertion of new or replacement of permanent pacemaker with transvenous electrode(s); atrial and ventricular |
| [33210](applewebdata://73F254D4-0729-44C2-A445-B1B00C8F2A86/../../../Downloads/%252525250a%2525252509%2525252509%2525252509%2525252509%2525252509%2525252509%2525252509%2525252509%2525252509Detail.aspx%2525253FCode=33210) | Insertion or replacement of temporary transvenous single chamber cardiac electrode or pacemaker catheter (separate procedure) |
| [33211](applewebdata://73F254D4-0729-44C2-A445-B1B00C8F2A86/../../../Downloads/%252525250a%2525252509%2525252509%2525252509%2525252509%2525252509%2525252509%2525252509%2525252509%2525252509Detail.aspx%2525253FCode=33211) | Insertion or replacement of temporary transvenous dual chamber pacing electrodes (separate procedure) |
| [33212](applewebdata://73F254D4-0729-44C2-A445-B1B00C8F2A86/../../../Downloads/%252525250a%2525252509%2525252509%2525252509%2525252509%2525252509%2525252509%2525252509%2525252509%2525252509Detail.aspx%2525253FCode=33212) | Insertion of pacemaker pulse generator only; with existing single lead |
| [33213](applewebdata://73F254D4-0729-44C2-A445-B1B00C8F2A86/../../../Downloads/%252525250a%2525252509%2525252509%2525252509%2525252509%2525252509%2525252509%2525252509%2525252509%2525252509Detail.aspx%2525253FCode=33213) | Insertion of pacemaker pulse generator only; with existing dual leads |
| [33214](applewebdata://73F254D4-0729-44C2-A445-B1B00C8F2A86/../../../Downloads/%252525250a%2525252509%2525252509%2525252509%2525252509%2525252509%2525252509%2525252509%2525252509%2525252509Detail.aspx%2525253FCode=33214) | Upgrade of implanted pacemaker system, conversion of single chamber system to dual chamber system (includes removal of previously placed pulse generator, testing of existing lead, insertion of new lead, insertion of new pulse generator) |
| [33215](applewebdata://73F254D4-0729-44C2-A445-B1B00C8F2A86/../../../Downloads/%252525250a%2525252509%2525252509%2525252509%2525252509%2525252509%2525252509%2525252509%2525252509%2525252509Detail.aspx%2525253FCode=33215) | Repositioning of previously implanted transvenous pacemaker or implantable defibrillator (right atrial or right ventricular) electrode |
| [33221](applewebdata://73F254D4-0729-44C2-A445-B1B00C8F2A86/../../../Downloads/%252525250a%2525252509%2525252509%2525252509%2525252509%2525252509%2525252509%2525252509%2525252509%2525252509Detail.aspx%2525253FCode=33221) | Insertion of pacemaker pulse generator only; with existing multiple leads |
| [33222](applewebdata://73F254D4-0729-44C2-A445-B1B00C8F2A86/../../../Downloads/%252525250a%2525252509%2525252509%2525252509%2525252509%2525252509%2525252509%2525252509%2525252509%2525252509Detail.aspx%2525253FCode=33222) | Relocation of skin pocket for pacemaker |
| [33224](applewebdata://73F254D4-0729-44C2-A445-B1B00C8F2A86/../../../Downloads/%252525250a%2525252509%2525252509%2525252509%2525252509%2525252509%2525252509%2525252509%2525252509%2525252509Detail.aspx%2525253FCode=33224) | Insertion of pacing electrode, cardiac venous system, for left ventricular pacing, with attachment to previously placed pacemaker or implantable defibrillator pulse generator (including revision of pocket, removal, insertion, and/or replacement of existing generator) |
| [33225](applewebdata://73F254D4-0729-44C2-A445-B1B00C8F2A86/../../../Downloads/%252525250a%2525252509%2525252509%2525252509%2525252509%2525252509%2525252509%2525252509%2525252509%2525252509Detail.aspx%2525253FCode=33225) | Insertion of pacing electrode, cardiac venous system, for left ventricular pacing, at time of insertion of implantable defibrillator or pacemaker pulse generator (eg, for upgrade to dual chamber system) (List separately in addition to code for primary procedure) |
| [33226](applewebdata://73F254D4-0729-44C2-A445-B1B00C8F2A86/../../../Downloads/%252525250a%2525252509%2525252509%2525252509%2525252509%2525252509%2525252509%2525252509%2525252509%2525252509Detail.aspx%2525253FCode=33226) | Repositioning of previously implanted cardiac venous system (left ventricular) electrode (including removal, insertion and/or replacement of existing generator) |
| [33227](applewebdata://73F254D4-0729-44C2-A445-B1B00C8F2A86/../../../Downloads/%252525250a%2525252509%2525252509%2525252509%2525252509%2525252509%2525252509%2525252509%2525252509%2525252509Detail.aspx%2525253FCode=33227) | Removal of permanent pacemaker pulse generator with replacement of pacemaker pulse generator; single lead system |
| [33228](applewebdata://73F254D4-0729-44C2-A445-B1B00C8F2A86/../../../Downloads/%252525250a%2525252509%2525252509%2525252509%2525252509%2525252509%2525252509%2525252509%2525252509%2525252509Detail.aspx%2525253FCode=33228) | Removal of permanent pacemaker pulse generator with replacement of pacemaker pulse generator; dual lead system |
| [33229](applewebdata://73F254D4-0729-44C2-A445-B1B00C8F2A86/../../../Downloads/%252525250a%2525252509%2525252509%2525252509%2525252509%2525252509%2525252509%2525252509%2525252509%2525252509Detail.aspx%2525253FCode=33229) | Removal of permanent pacemaker pulse generator with replacement of pacemaker pulse generator; multiple lead system |
| [33233](applewebdata://73F254D4-0729-44C2-A445-B1B00C8F2A86/../../../Downloads/%252525250a%2525252509%2525252509%2525252509%2525252509%2525252509%2525252509%2525252509%2525252509%2525252509Detail.aspx%2525253FCode=33233) | Removal of permanent pacemaker pulse generator only |
| [33234](applewebdata://73F254D4-0729-44C2-A445-B1B00C8F2A86/../../../Downloads/%252525250a%2525252509%2525252509%2525252509%2525252509%2525252509%2525252509%2525252509%2525252509%2525252509Detail.aspx%2525253FCode=33234) | Removal of transvenous pacemaker electrode(s); single lead system, atrial or ventricular |
| [33235](applewebdata://73F254D4-0729-44C2-A445-B1B00C8F2A86/../../../Downloads/%252525250a%2525252509%2525252509%2525252509%2525252509%2525252509%2525252509%2525252509%2525252509%2525252509Detail.aspx%2525253FCode=33235) | Removal of transvenous pacemaker electrode(s); dual lead system |
| [33236](applewebdata://73F254D4-0729-44C2-A445-B1B00C8F2A86/../../../Downloads/%252525250a%2525252509%2525252509%2525252509%2525252509%2525252509%2525252509%2525252509%2525252509%2525252509Detail.aspx%2525253FCode=33236) | Removal of permanent epicardial pacemaker and electrodes by thoracotomy; single lead system, atrial or ventricular |
| [33237](applewebdata://73F254D4-0729-44C2-A445-B1B00C8F2A86/../../../Downloads/%252525250a%2525252509%2525252509%2525252509%2525252509%2525252509%2525252509%2525252509%2525252509%2525252509Detail.aspx%2525253FCode=33237) | Removal of permanent epicardial pacemaker and electrodes by thoracotomy; dual lead system |
| [33238](applewebdata://73F254D4-0729-44C2-A445-B1B00C8F2A86/../../../Downloads/%252525250a%2525252509%2525252509%2525252509%2525252509%2525252509%2525252509%2525252509%2525252509%2525252509Detail.aspx%2525253FCode=33238) | Removal of permanent transvenous electrode(s) by thoracotomy |
| [33274](applewebdata://73F254D4-0729-44C2-A445-B1B00C8F2A86/../../../Downloads/%252525250a%2525252509%2525252509%2525252509%2525252509%2525252509%2525252509%2525252509%2525252509%2525252509Detail.aspx%2525253FCode=33274) | Transcatheter insertion or replacement of permanent leadless pacemaker, right ventricular, including imaging guidance (eg, fluoroscopy, venous ultrasound, ventriculography, femoral venography) and device evaluation (eg, interrogation or programming), when performed |
| [33275](applewebdata://73F254D4-0729-44C2-A445-B1B00C8F2A86/../../../Downloads/%252525250a%2525252509%2525252509%2525252509%2525252509%2525252509%2525252509%2525252509%2525252509%2525252509Detail.aspx%2525253FCode=33275) | Transcatheter removal of permanent leadless pacemaker, right ventricular |
| 71090 | Insertion pacemaker, fluoroscopy and radiography, radiological supervision and interpretation |
| [93279](applewebdata://73F254D4-0729-44C2-A445-B1B00C8F2A86/../../../Downloads/%252525250a%2525252509%2525252509%2525252509%2525252509%2525252509%2525252509%2525252509%2525252509%2525252509Detail.aspx%2525253FCode=93279) | Programming device evaluation (in person) with iterative adjustment of the implantable device to test the function of the device and select optimal permanent programmed values with analysis, review and report by a physician or other qualified health care professional; single lead pacemaker system or leadless pacemaker system in one cardiac chamber |
| [93280](applewebdata://73F254D4-0729-44C2-A445-B1B00C8F2A86/../../../Downloads/%252525250a%2525252509%2525252509%2525252509%2525252509%2525252509%2525252509%2525252509%2525252509%2525252509Detail.aspx%2525253FCode=93280) | Programming device evaluation (in person) with iterative adjustment of the implantable device to test the function of the device and select optimal permanent programmed values with analysis, review and report by a physician or other qualified health care professional; dual lead pacemaker system |
| [93281](applewebdata://73F254D4-0729-44C2-A445-B1B00C8F2A86/../../../Downloads/%252525250a%2525252509%2525252509%2525252509%2525252509%2525252509%2525252509%2525252509%2525252509%2525252509Detail.aspx%2525253FCode=93281) | Programming device evaluation (in person) with iterative adjustment of the implantable device to test the function of the device and select optimal permanent programmed values with analysis, review and report by a physician or other qualified health care professional; multiple lead pacemaker system |
| [93286](applewebdata://73F254D4-0729-44C2-A445-B1B00C8F2A86/../../../Downloads/%252525250a%2525252509%2525252509%2525252509%2525252509%2525252509%2525252509%2525252509%2525252509%2525252509Detail.aspx%2525253FCode=93286) | Peri-procedural device evaluation (in person) and programming of device system parameters before or after a surgery, procedure, or test with analysis, review and report by a physician or other qualified health care professional; single, dual, or multiple lead pacemaker system, or leadless pacemaker system |
| [93288](applewebdata://73F254D4-0729-44C2-A445-B1B00C8F2A86/../../../Downloads/%252525250a%2525252509%2525252509%2525252509%2525252509%2525252509%2525252509%2525252509%2525252509%2525252509Detail.aspx%2525253FCode=93288) | Interrogation device evaluation (in person) with analysis, review and report by a physician or other qualified health care professional, includes connection, recording and disconnection per patient encounter; single, dual, or multiple lead pacemaker system, or leadless pacemaker system |
| [93293](applewebdata://73F254D4-0729-44C2-A445-B1B00C8F2A86/../../../Downloads/%252525250a%2525252509%2525252509%2525252509%2525252509%2525252509%2525252509%2525252509%2525252509%2525252509Detail.aspx%2525253FCode=93293) | Transtelephonic rhythm strip pacemaker evaluation(s) single, dual, or multiple lead pacemaker system, includes recording with and without magnet application with analysis, review and report(s) by a physician or other qualified health care professional, up to 90 days |
| [93294](applewebdata://73F254D4-0729-44C2-A445-B1B00C8F2A86/../../../Downloads/%252525250a%2525252509%2525252509%2525252509%2525252509%2525252509%2525252509%2525252509%2525252509%2525252509Detail.aspx%2525253FCode=93294) | Interrogation device evaluation(s) (remote), up to 90 days; single, dual, or multiple lead pacemaker system, or leadless pacemaker system with interim analysis, review(s) and report(s) by a physician or other qualified health care professional |
| [93296](applewebdata://73F254D4-0729-44C2-A445-B1B00C8F2A86/../../../Downloads/%252525250a%2525252509%2525252509%2525252509%2525252509%2525252509%2525252509%2525252509%2525252509%2525252509Detail.aspx%2525253FCode=93296) | Interrogation device evaluation(s) (remote), up to 90 days; single, dual, or multiple lead pacemaker system, leadless pacemaker system, or implantable defibrillator system, remote data acquisition(s), receipt of transmissions and technician review, technical support and distribution of results |
| 93731 | Analyze pacemaker system |
| 93732 | Analyze pacemaker system |
| 93733 | Telephone analysis, pacemaker |
| 93734 | Analyze pacemaker system |
| 93735 | Analyze pacemaker system |
| 93736 | Telephonic analysis pacemaker |
